# Striatal dopamine depletion drives disease progression and network topology aberrations specifically by impairing left M1 network

**DOI:** 10.1101/2023.09.19.23295781

**Authors:** Zhichun Chen, Guanglu Li, Liche Zhou, Lina Zhang, Jun Liu

## Abstract

**Background:** Stratal dopamine depletion contributes to both motor and non-motor symptoms of patients with Parkinson’s disease (PD). The objective of current study is to explore whether stratal dopamine depletion shapes clinical heterogeneity by impairing brain networks of PD patients.

**Methods:** In this cross-sectional study, PD participants undergoing functional magnetic resonance imaging from Parkinson’s Progression Markers Initiative (PPMI) database were investigated. According to the levels of striatal binding ratio (SBR) in bilateral striatum, PD patients were classified into lower quartile group (SBR level rank: 0%∼25%), interquartile group (SBR level rank: 26%∼75%), and upper quartile group (SBR level rank: 76%∼100%) based on their SBR level quartiles to examine how stratal dopamine depletion affects clinical manifestations and brain networks.

**Findings:** PD patients in the lower quartile group showed more severe motor and non-motor symptoms compared to upper quartile group. Additionally, topological metrics in both structural and functional network were significantly different between upper quartile group and lower quartile group. Furthermore, the functional network of left primary motor cortex (M1) was specifically impaired in lower quartile group, which resulted in topological disruptions in functional network. Importantly, impaired left M1 network in PD patients mediated the effects of striatal dopamine depletion on both motor and non-motor symptoms.

**Interpretation:** Striatal dopamine depletion specifically impaired left M1 network, which contributed to aberrant functional network topology and dopamine-dependent motor and non-motor symptoms.

**Funding:** National Natural Science Foundation of China (Grant No. 81873778, 82071415) and National Research Center for Translational Medicine at Shanghai (Grant No. NRCTM(SH)-2021-03).

## Introduction

Parkinson’s disease (PD) is the most widespread movement disorder and the second most prevalent neurodegenerative diseases, trailing behind Alzheimer’s disease (AD).^1,2^ In addition to the classical motor symptoms, which include bradykinesia, resting tremor, and rigidity, PD is also linked to a diverse range of non-motor symptoms that significantly impact the overall burden of the disease, including REM sleep behavior disorder (RBD), hypersomnia, autonomic dysfunction, depression, anxiety, apathy, and cognitive impairment.^1,2^ One of the most prominent pathological hallmarks of PD is the loss of dopaminergic neurons in the substantia nigra,^3^ resulting in the deficiency of dopamine in the striatum.^4^ Dopamine transporter imaging using single-photon emission computed tomography (DAT-SPECT) is one of the most highly developed supplementary assessments for PD, which is utilized to indirectly measure DAT availability in straitum.^5,6^ Striatal binding ratio (SBR) in striatum measured by DAT-SPECT has been demonstrated to be associated with motor manifestations, including rigidity,^7,8^ resting tremor,^9^ and bradykinesia.^8^ Apart from its associations with motor symptoms, striatum SBR was also linked to multiple non-motor symptoms, such as RBD,^10^ cognitive impairment,^10–13^ anxiety,^10^ apathy,^14^ depression,^15,16^ impulse control disorders,^17^ sexual dysfunction,^18^ urinary dysfunction,^19^ cardiovascular dysfunction,^20^ gastrointestinal dysfunction,^21^ and olfaction loss.^22^ Taken together, striatum SBR is an essential indicator of dopaminergic neurodegeneration in PD, which intimately correlated with both motor and non-motor symptoms.

Dopamine is a key neurotransmitter involved in the regulation of neural activity ^23,24^ and modulates blood oxygenation level-dependent (BOLD) signals obtained by functional magnetic resonance imaging (fMRI).^25,26^ Dopamine also regulates functional connectivity (FC) and functional networks measured by fMRI.^27–29^ Striatal dopamine depletion is associated with widespread alterations of brain functional networks in PD ^30,31^ while dopamine replacement normalizes aberrations of functional networks in PD.^32^ In addition, striatal dopamine depletion is also associated with significant changes of white matter network ^33^ and gray matter structure ^34^ in PD. Therefore, striatal dopamine depletion is significantly associated with both functional and structural networks in PD. Given that both brain structural and functional networks were significantly associated with motor and non-motor symptoms in PD,^35–37^ it is highly possible that brain network alterations may mediate the associations between striatum SBR and motor and non-motor manifestations of PD patients.

As mentioned above, striatum SBR is associated with both motor and non-motor symptoms in PD, however, the neural mechanisms underlying the associations between striatum SBR and clinical features of PD patients are poorly understood. Recently, we reported topological properties of brain networks were associated with both motor and non-motor symptoms of PD patients.^35,38,39^ In this study, we hypothesize that striatum SBR may shape motor and non-motor symptoms by modulating functional and structural networks of PD patients. As a consequence, the objective of this study is to examine whether structural and functional networks mediate the effects of striatum SBR on clinical features of PD patients. Specifically, the goals of this study include: (i) to validate the associations between striatum SBR and clinical features revealed by previous studies; (ii) to evaluate the effects of striatum SBR on structural and functional networks; (iii) to examine the associations between clinical features and imaging metrics; (iv) to investigate whether brain networks mediate the effects of striatum SBR on clinical features of PD patients with mediation analysis.

## Methods

### Participants

The data of PD participants from the Parkinson Progression Markers Initiative (PPMI) study ^40,41^ are downloaded and investigated. The Institutional Review Committee of all participating sites approved the PPMI study. The informed consents have been signed by all study participants, which can be obtained from the site investigators. The inclusion criteria for PD patients were shown as below: (i) The participants were over the age of 30; (ii) The participants were diagnosed with PD based on the MDS Clinical Diagnostic Criteria for PD; (iii) The participants underwent 3D T1-weighted MPRAGE imaging, resting-state fMRI, and diffusion tensor imaging (DTI) simultaneously. The exclusion criteria for PD patients were as follows: (i) The participants had significant abnormalities in the T1-weighted and T2-weighted MRI; (ii) The participants had genetic mutations associated with familial PD or were from the genetic PPMI cohort and prodromal cohort. According to above inclusion and exclusion criteria, totally 141 PD participants were included for the final analysis. The motor symptoms of PD patients were evaluated, including Hoehn and Yahr (H&Y) stages, Total Rigidity scores, Tremor scores, and the MDS Unified Parkinson’s Disease Rating Scale (MDS-UPDRS). The non-motor symptoms were assessed with Epworth Sleepiness Scale (ESS), REM Sleep Behavior Disorder Screening Questionnaire (RBDSQ), Scale for Outcomes in Parkinson’s Disease-Autonomic (SCOPA-AUT), 15-item Geriatric Depression Scale (GDS), Benton Judgment of Line Orientation Test (BJLOT), Letter Number Sequencing Test (LNS), Semantic Fluency Test (SFT), Symbol Digit Modalities Test (SDMT), and Montreal Cognitive Assessment (MoCA). The patients also obtained [^123^I] FP-CIT SPECT scans, which were examined in accordance with the technical manual of the PPMI study (http://ppmi-info.org/). The SBRs for bilateral caudate, putamen, and striatum were derived from SPECT scans. They were calculated with the formula: (target region/reference region)-1, in which occipital lobe was the reference region. To examine the effects of striatum SBR on clinical assessments and brain networks, PD patients (n = 141) were divided into lower quartile group (Q1, SBR level rank: 0%∼25%, SBR range: 0.40∼0.99), interquartile group (Q2-3, SBR level rank: 26%∼75%, SBR range: 0.99∼1.52), and upper quartile group (Q4, SBR level rank: 76%∼100%, SBR range: 1.53∼2.73) based on their SBR level quartiles in bilateral striatum. The clinical features of participants in each group were shown in Table S1 and Figure 1. The similar approaches for subgroup divisions based on quartiles of continuous variables have been described previously.^35,42,43^

**Figure 1.**
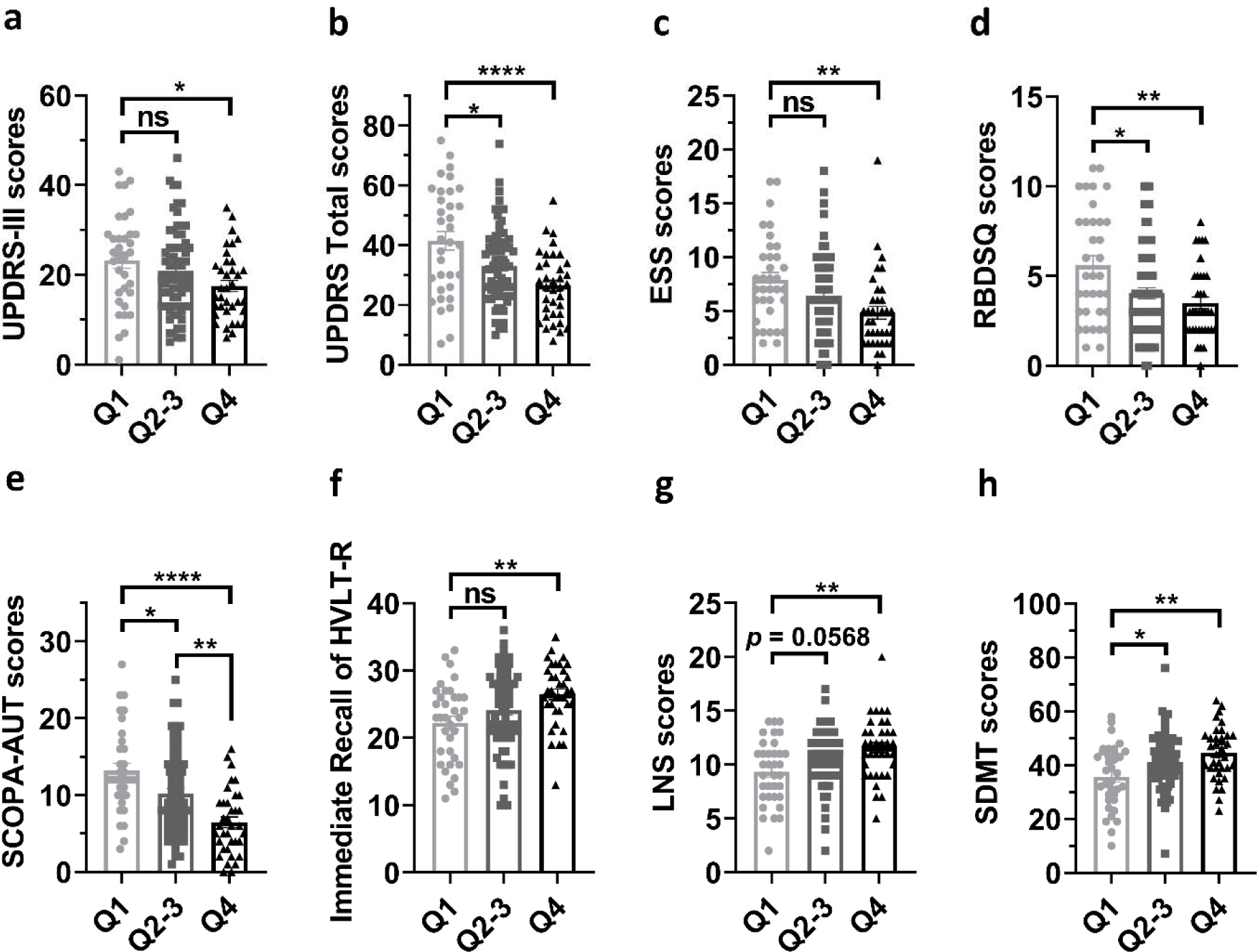
Group differences of clinical characteristics. (a-h) Group differences of scores of UPDRS-III (a), total UPDRS (b), ESS (c), RBDSQ (d), SCOPA-AUT (e), immediate recall of HVLT-R (f), LNS (g), and SDMT (h). One-way ANOVA followed by Tukey’s post hoc test (Q1 group *vs* Q2-3 group *vs* Q4 group) were conducted to compare clinical variables. *p* < 0.05 was considered statistically significant. \**p* < 0.05, \*\**p* < 0.01, \*\*\*\**p* < 0.0001. Abbreviations: UPDRS, Unified Parkinson’s Disease Rating Scale; ESS, Epworth Sleepiness Scale; RBDSQ, REM Sleep Behavior Disorder Screening Questionnaire; SCOPA-AUT, Scale for Outcomes in Parkinson’s Disease-Autonomic; HVLT-R, Hopkins Verbal Learning Test–Revised; LNS, Letter Number Sequencing test; SDMT, Symbol Digit Modalities Test.

### Image acquisition

The T1-weighted MRI images in three dimensions were obtained using 3T Siemens (TIM Trio or Verio) scanners from Siemens Healthcare, utilizing a magnetization-prepared rapid acquisition gradient echo sequence. The parameters for T1 images were as follows: Repetition time (TR) = 2300 ms, Echo time (TE) = 2.98 ms, Voxel size = 1 mm^3^, Slice thickness = 1.2 mm, twofold acceleration, and sagittal-oblique angulation. The DTI was conducted with the following parameters: TR = 8,400-8,800 ms, TE = 88 ms, Voxel size = 2 mm^3^, Slice thickness = 2 mm, 64 directions, b-value = 1000 s/mm^2^. The resting-state fMRI was acquired using a T2*-weighted echo planar imaging (EPI) sequence with the following parameters: TR = 2400 ms, TE = 25 ms, Voxel size = 3.3 mm^3^, Slice number = 40, Slice thickness = 3.3 mm, Flip angle = 90°, and Acquisition time = 8 min.

### Imaging preprocessing

DTI images of 141 PD participants were preprocessed using the FMRIB Software Library toolkit (FSL, https://fsl.fmrib.ox.ac.uk/fsl/fslwiki). Briefly, the DTI images initially underwent corrections for head motions, distortions caused by eddy currents, and susceptibility artifacts resulting from magnetic field inhomogeneity. Then, DTI metrics, such as fractional anisotropy (FA), mean diffusivity (MD), axial diffusivity, and radial diffusivity, were generated. Subsequently, the individual processed images were further reconstructed in the standard MNI space for participant comparisons.

Among 141 PD patients with DTI images, only 82 of them obtained the resting-state fMRI images. The preprocessing of resting-state fMRI images was carried out using SPM12 (http://www.fil.ion.ucl.ac.uk/spm/software/spm12/) and GRETNA software (https://www.nitrc.org/projects/gretna/). As demonstrated by our recent studies, the preprocessing steps included standard procedures for correcting slice-timing, normalizing spatially using a standard EPI template, smoothing spatially (4-mm Gaussian kernel), eliminating nuisance signals (head motion profiles, CSF signal, white matter signals, and local and global hardware artifacts), and filtering temporally within the frequency range of 0.01–0.1 Hz. Based on previous investigations, we excluded 8 participants who had head motion frame-wise displacement (FD) greater than 0.5 mm and head rotation exceeding 2° due to head movements ^44–47^. According to our recent study, the demographic and clinical characteristics were not significantly different between PD patients with DTI images and PD patients with preprocessed resting-state images (n = 74).^38^ Furthermore, we adjusted the effects of head motions on the functional images by regressing out the head motion parameters. Ultimately, we revealed no significant differences in FD values among three quartile groups (*p* > 0.05).

### Network construction

A complimentary MATLAB toolkit, PANDA (http://www.nitrc.org/projects/panda/), was employed to execute deterministic fiber tractography to construct the white matter network. The Fiber Assignment by Continuous Tracking (FACT) algorithm was utilized to compute whole-brain white matter fibers between each pair of 90 cortical and subcortical nodes in the Automated Anatomical Labeling (AAL) atlas. The threshold for FA skeleton was set at 0.20 and a threshold of 45° was set for fiber angle. After the white matter tractography, a white matter network matrix based on fiber number (FN) was generated for each participant.

The construction of functional network matrix was carried out in the following manner. Initially, ninety cortical and subcortical nodes were established using the AAL atlas. Subsequently, the pairwise FC was computed as the Pearson correlation coefficient of the time series of 90 nodes. The Fisher’s r-to-z conversions of correlation coefficients were executed to enhance the normality of FC. Ultimately, a 90 x 90 functional matrix was generated for each participant.

### Graph-based network analysis

The graphical network measurements of brain networks were computed using GRETNA (https://www.nitrc.org/projects/gretna/). A range of network density thresholds (0.05 ∼ 0.50 with a step size of 0.05) was utilized to compute the global and nodal network measurements. The area under curve (AUC) of global and nodal network measurements was calculated. The global network properties consisted of global efficiency, local efficiency, and small-worldness measurements (clustering coefficient [Cp], characteristic path length [Lp], normalized clustering coefficient [γ], normalized characteristic path length [λ], and small worldness [σ]). The nodal network properties consisted of: nodal betweenness centrality (BC), nodal degree centrality (DC), nodal Cp, nodal efficiency, nodal local efficiency, and nodal shortest path length. The detailed definitions for each network measurement have been documented by previous studies.^38,48,49^

## Statistical analysis

### Comparison of clinical variables and multivariate regression analysis

One-way ANOVA followed by Tukey’s post hoc test (Q1 group *vs* Q2-3 group *vs* Q4 group) was employed to compare continuous clinical variables. χ^2^ test was employed to compare categorical variables. The multivariate regression analysis with age, sex, disease duration, and years of education as covariates was used to analyze the associations between striatum SBR and clinical variables. *p* < 0.05 was considered statistically significant.

### Comparisons of network metrics and multivariate regression analysis

For comparisons of network measurements, two-way ANOVA test was utilized. The areas under the curve (AUCs) of global network measurements were compared using one-way ANOVA test. The multivariate regression analysis with age, sex, disease duration, and years of education as covariates was used to analyze the associations between striatum SBR and network metrics. False discovery rate (FDR)-corrected *p* < 0.05 was considered to be statistically significant.

### Comparison of global network strengths and node-based FCs

Network-Based Statistic (NBS, https://www.nitrc.org/projects/nbs/) method was utilized to compare the global network strengths of brain networks among 3 quartile groups. Age, sex, years of education, and disease duration were included as covariates during NBS analysis. The FC of left M1 was compared among 3 quartile groups using two-way ANOVA test. *p* < 0.05 after FDR correction was considered statistically significant.

### Construction of correlation matrix for imaging metrics

The matrices for graphical metric autocorrelations or correlations between graphical metrics and FC of left M1 were constructed with Pearson’s correlation method. FDR-corrected *p* < 0.05 was considered to be statistically significant.

### Association analysis between imaging metrics and clinical variables

The association analysis between graphical metrics and clinical variables or between FC of left M1 and clinical variables was performed with both Pearson’s correlation and multivariate regression analysis. During multivariate regression analysis, age, sex, years of education, and disease duration were included as covariates. FDR-corrected *p* < 0.05 was considered to be statistically significant.

### Mediation analysis

IBM SPSS Statistics Version 26 was employed to conduct mediation analysis. The independent variable in the mediation model was the SBR in bilateral putamen or striatum. The dependent variable was scores of clinical assessments (UPDRS-III, total UPDRS, SDMT, RBDSQ, and SCOPA-AUT) or graphical network metrics. The mediators were topological network metrics or FC of left M1. We modeled the indirect effects of mediators on the relationships between striatum SBR and clinical assessments or topological network metrics. Age, sex, disease duration, and years of education were included as confounding variables during the mediation analysis. *p* < 0.05 was considered statistically significant.

## Results

### The effects of striatum SBR on clinical assessments

The demographic data and clinical characteristics of participants in each group were compared and shown in Table S1 and Figure 1.

### The associations between striatum SBR and clinical variables

In 141 PD patients, after adjusting age, sex, years of education, and disease duration, striatum SBR was significantly associated with both motor and non-motor assessments (Table 1).

**Table 1:** Associations between DAT availability and clinical assessments.

| Clinical assessments |  |  |  |  |  |  |  |  |
| --- | --- | --- | --- | --- | --- | --- | --- | --- |
|  | HY stage | Tremor scores | Rigidity scores | UPDRS-III scores | UPDRS Total scores | ESS scores |  |  |
| $\beta$ value | $\beta = -0.20$ | $\beta = 2.27$ | $\beta = -1.37$ | $\beta = -4.20$ | $\beta = -11.64$ | $\beta = -2.44$ | | |
| $p$-value | $p = 0.0808$ | $p = \mathbf{0.0024}$ | $p = \mathbf{0.0385}$ | $p = \mathbf{0.0553}$ | $p = \mathbf{0.0005}$ | $p = \mathbf{0.0064}$ | | |
| Clinical assessments |  |  |  |  |  |  |  |  |
|  | RBDSQ scores | SCOPA-AUT scores | LNS scores | BJLOT scores | SFT scores | SDMT scores | MoCA scores | HVLT-R Immediate Recall |
| $\beta$ value | $\beta = -1.43$ | $\beta = -4.98$ | $\beta = 1.56$ | $\beta = 0.46$ | $\beta = 1.65$ | $\beta = 5.80$ | $\beta = 0.47$ | $\beta = 1.88$ |
| $p$-value | $p = \mathbf{0.0214}$ | $p < \mathbf{0.0001}$ | $p = \mathbf{0.0073}$ | $p = 0.2913$ | $p = 0.5020$ | $p = \mathbf{0.0095}$ | $p = 0.4292$ | $p = 0.1201$ |

The data were shown as the $\beta$ and $p$ -values derived from multivariate regression analysis with age, sex, years of education, and disease duration as covariates. The motor function examination was assessed in ON state. $p < 0.05$ was considered statistically significant. The bold values indicate statistical significance. Abbreviations: HY, Hoehn & Yahr; UPDRS, Unified Parkinson's Disease Rating Scale; ESS, Epworth Sleepiness Scale; RBDSQ, REM Sleep Behavior Disorder Screening Questionnaire; SCOPA-AUT, Scale for Outcomes in Parkinson's Disease-Autonomic; SDMT, Symbol Digit Modalities Test; LNS, Letter Number Sequencing; SFT, Semantic Fluency Test Score; BJLOT, Benton Judgement of Line Orientation; MoCA, Montreal Cognitive Assessment; HVLT-R, Hopkins Verbal Learning Test-Revised.

### The effects of striatum SBR on the topology of brain networks

The global network metrics in both functional network and structural network were not statistically different among 3 quartile groups (all *p* > 0.05 after FDR correction). In structural network, PD patients in Q1 group showed significant different nodal network metrics in multiple brain regions compared to Q4 group (FDR-corrected *p* < 0.05; Supplementary Figure 1). For functional network, nodal network metrics were also significantly different among 3 quartile groups (FDR-corrected *p* < 0.05; Figure 2a-f).

**Figure 2.**
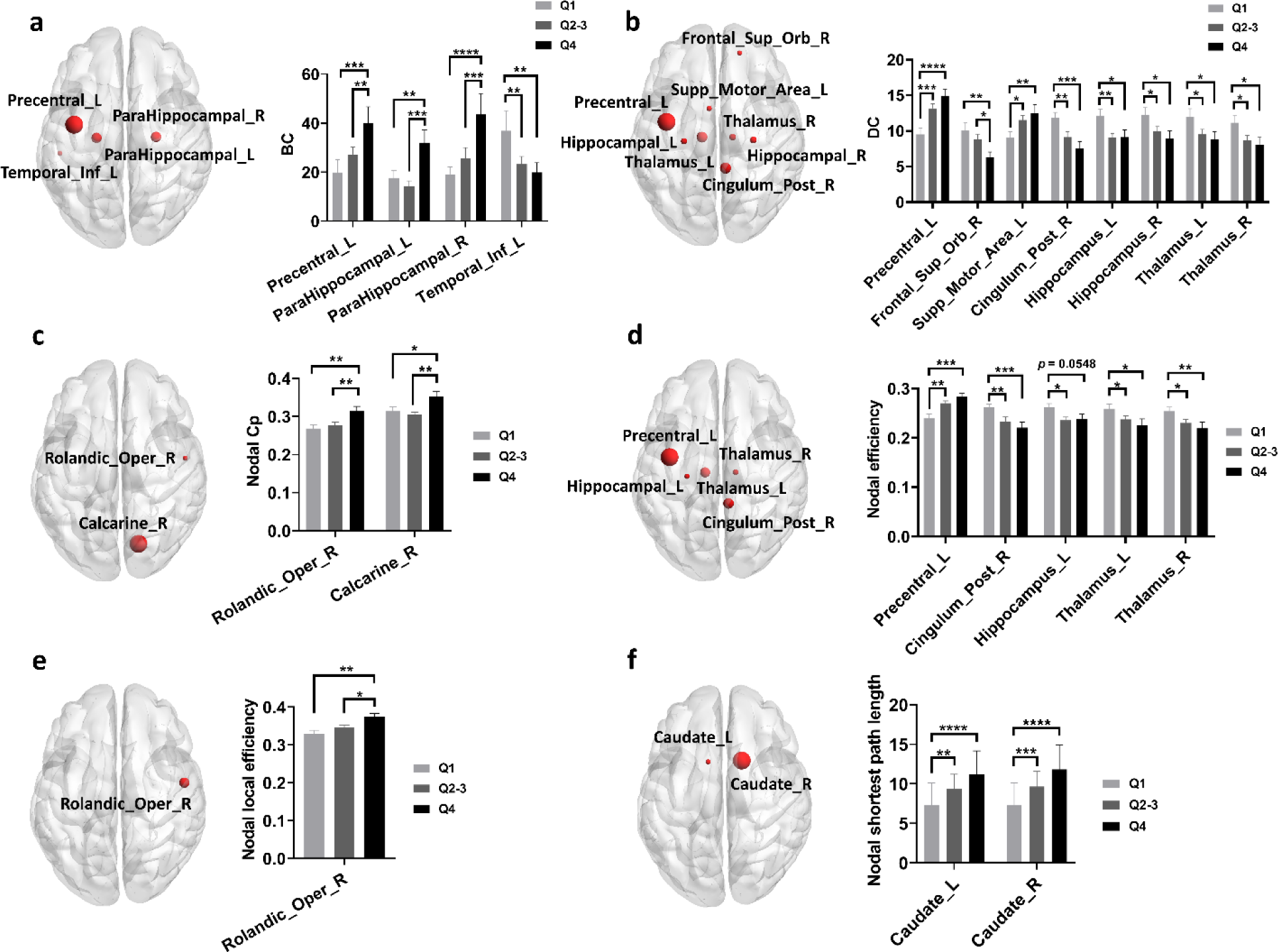
Group differences of nodal network metrics in functional network. (a-f) Group differences of nodal BC (a), nodal DC (b), nodal Cp (c), nodal efficiency (d), nodal local efficiency (e), and nodal shortest path length (f) in functional network among different quartile groups. Two-way ANOVA test with FDR correction was performed. *p* < 0.05 after FDR correction was considered to have statistical significance. \**p* < 0.05, ** *p* < 0.01, \*\*\**p* < 0.001, \*\*\*\**p* < 0.0001. Abbreviations: BC, Betweenness centrality; DC, Degree centrality; Cp, Clustering coefficient.

### Associations between striatum SBR and brain network metrics

As shown in Supplementary Figure 2, striatum SBR was associated with multiple structural network metrics (Supplementary Figure 2). In comparison with structural network metrics, striatum SBR showed much stronger correlations with functional network metrics. Specifically, striatum SBR was positively associated with multiple nodal network metrics in functional network (Supplementary Figure 3a-d). Similarly, caudate SBR and putamen SBR were also strongly and positively correlated with above functional network metrics (Supplementary Figure 3e-l).

### The effects of striatal dopamine depletion on global network strengths

As shown above, we revealed striatum SBR significantly shaped the topology of functional and structural network, therefore, we hypothesized that striatum SBR may affect network topology by modifying structural and functional connectivity strengths. With NBS analysis, no significant differences in global network strengths of structural network were observed among 3 quartile groups, however, we revealed significant differences in global network strengths of functional network between Q1 group and Q4 group (FDR-corrected *p* < 0.05, NBS analysis; Figure 3). Specifically, Q1 group exhibited significantly lower global network strengths in functional network compared to Q4 group, especially in a subnetwork with left precentral gyrus (M1, Primary motor cortex) as the core node.

**Figure 3.**
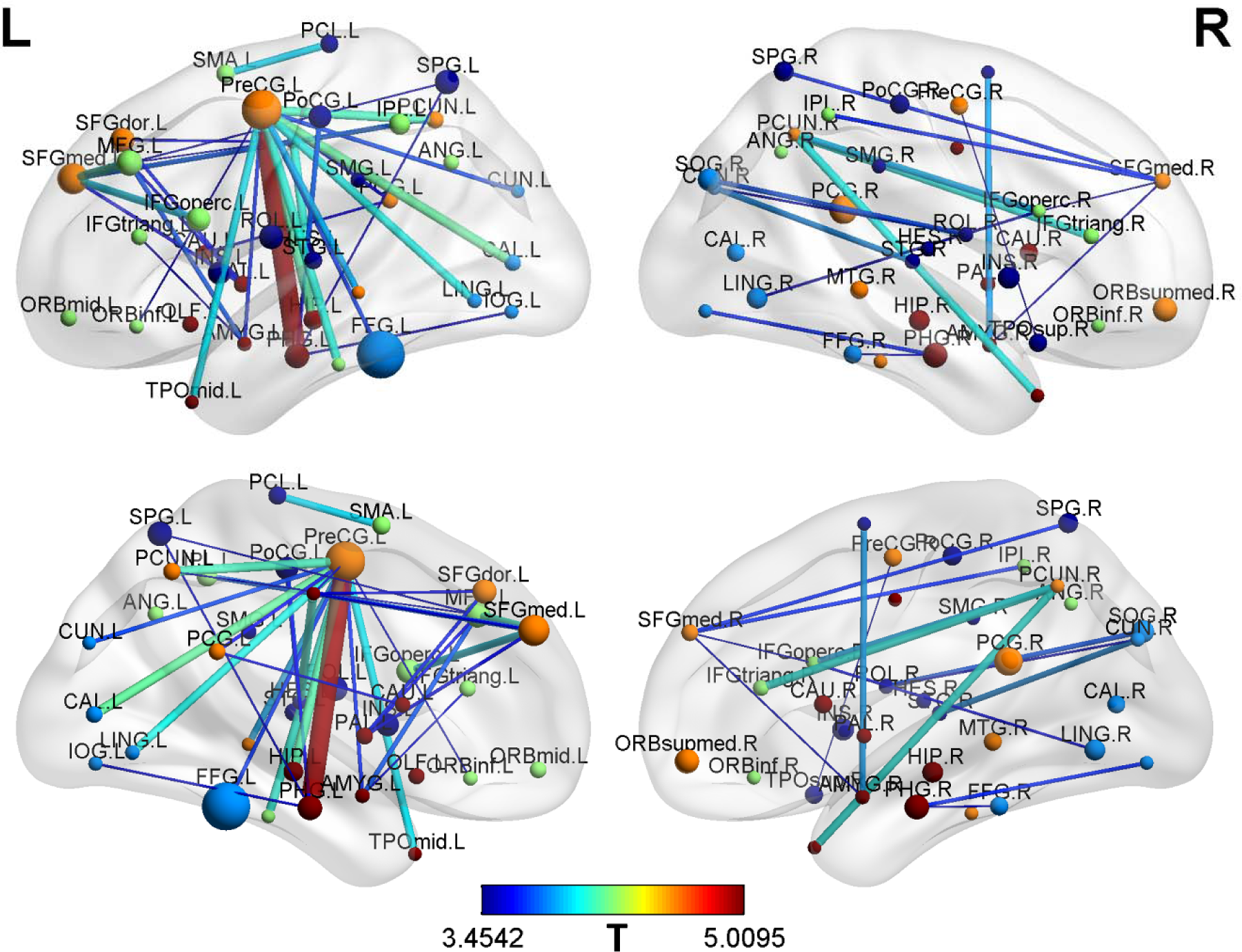
Lower global network strengths in Q1 group compared to Q4 group. Q1 group exhibited significantly lower global network strengths in functional network compared to Q4 group, especially in a subnetwork with left M1 (primary motor cortex) as core node. NBS analysis was utilized to compare the global network strength of brain networks among 3 quartile groups. FDR-corrected *p* < 0.05 was considered statistically significant. Age, sex, years of education, and disease duration, were included as covariates during NBS analysis.

### The effects of striatal dopamine depletion on FC of left M1

As shown in Figure 3, FCs between left M1 and multiple nodes were much lower in Q1 group compared to Q4 group. With node-based connectivity analysis, we further demonstrated that FCs between left M1 and a multitude of brain nodes in AAL atlas were much lower in Q1 group compared to Q4 group, spanning frontal, temporal, occipital, parietal, subcortical, and cingulate areas (FDR-corrected *p* < 0.05, Two-way ANOVA test; Figure 4a-f).

**Figure 4.**
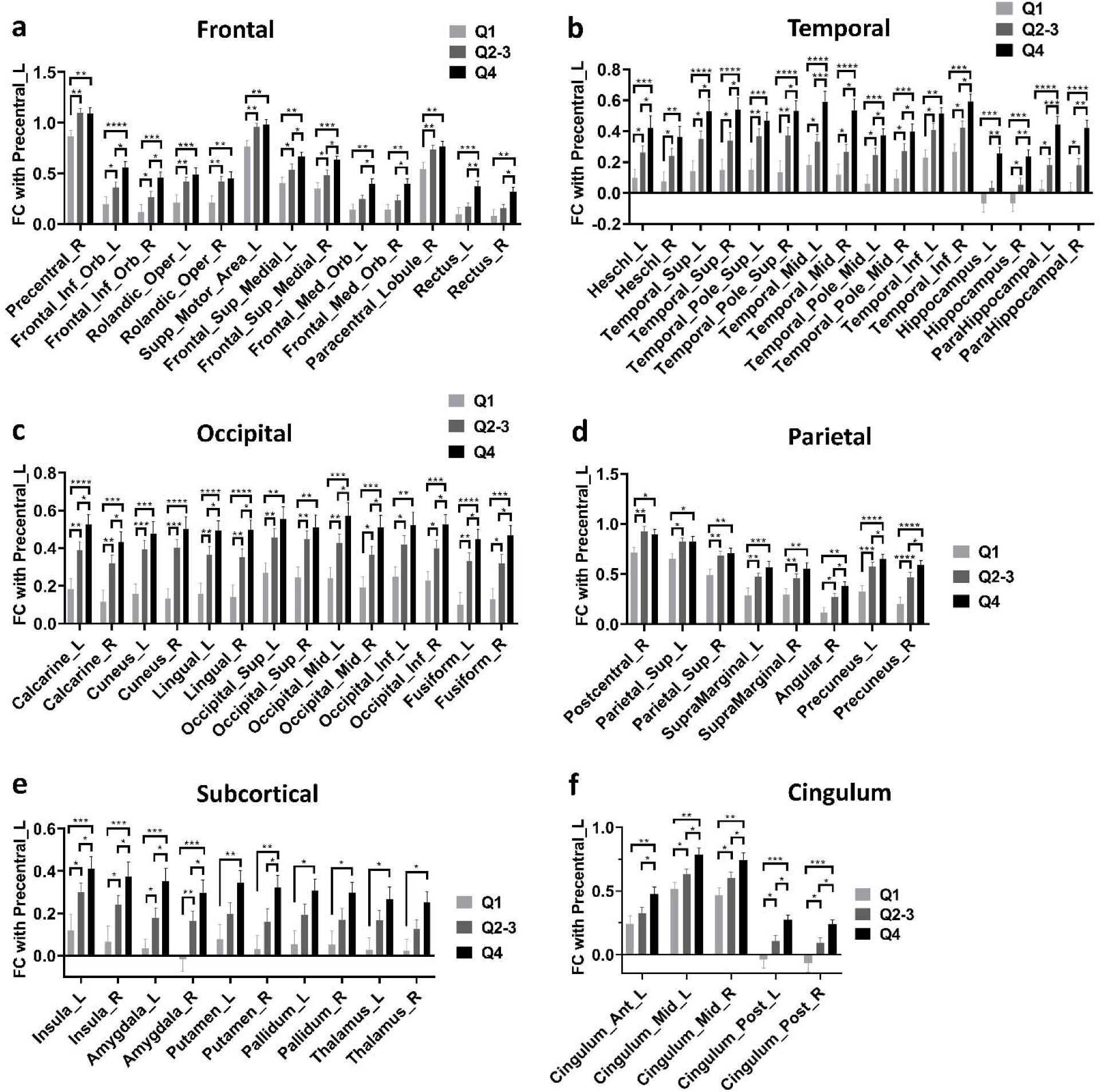
Lower FC of left M1 in Q1 group compared to Q4 group. (a-f) Q1 group exhibited lower FC between left M1 and a multitude of brain nodes compared to Q4 group, spanning frontal (a), temporal (b), occipital (c), parietal (d), subcortical (e), and cingulate areas (FDR-corrected *p* < 0.05, Two-way ANOVA test). Two-way ANOVA test with FDR correction was performed. *p* < 0.05 after FDR correction was considered to have statistical significance. \**p* < 0.05, ** *p* < 0.01, \*\*\**p* < 0.001, \*\*\*\**p* < 0.0001. Abbreviations: FC, Functional connectivity.

### The associations between FC of left M1 and topological metrics

As shown in Figure 5a, we initially found significant autocorrelations between nodal topological metrics showing statistical differences in Figure 2. Specifically, nodal DC and nodal efficiency in multiple nodes were auto-correlated and nodal DC were also significantly correlated with nodal efficiency in multiple nodes (FDR-corrected *p* < 0.05; Figure 5a). In addition, nodal DC in left M1 was substantially correlated with multiple nodal metrics (FDR-corrected *p* < 0.05; Figure 5a). As shown in Figure 5b, FCs between left M1 and multiple nodes were significantly correlated with altered topological metrics (FDR-corrected *p* < 0.05; Figure 5b). Specifically, FCs between left M1 and multiple nodes were significantly correlated with nodal BC, DC, and efficiency of left M1 (FDR-corrected *p* < 0.05; Figure 5b).

**Figure 5.**
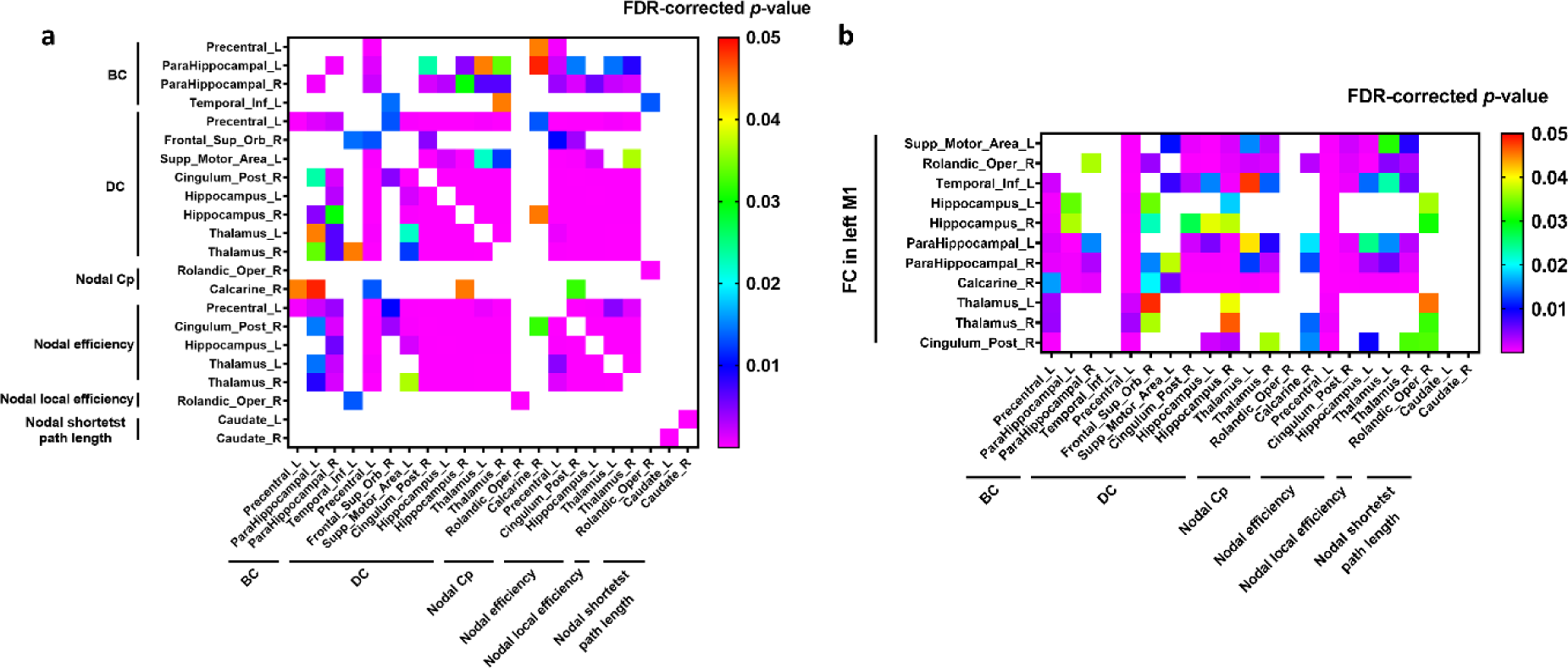
Significant correlations between FC of left M1 and functional topological metrics. (a) Functional topological metrics were autocorrelated. Specifically, nodal DC and nodal efficiency in multiple nodes were auto-correlated and nodal DC were also significantly correlated with nodal efficiency in multiple nodes (FDR-corrected *p* < 0.05). (b) FC between left M1 and multiple nodes were significantly correlated with altered functional topological metrics showing statistical significance in Figure 2 (FDR-corrected *p* < 0.05). The correlation analysis was conducted by Pearson correlation method. FDR-corrected *p* < 0.05 was considered statistically significant. Abbreviations: BC, Betweenness centrality; DC, Degree centrality; Cp, Clustering coefficient; FC, Functional connectivity; M1, Primary motor cortex.

To further examine whether alterations in FC of left M1 were causally associated with topological changes of functional network, we conducted mediation analysis to evaluate the causal associations. As shown in Figure 6, FC between left M1 and left inferior temporal gyrus, left hippocampus, right hippocampus, and right posterior cingulate cortex mediated the effects of striatum SBR on nodal BC, DC, and efficiency of left M1 (FDR-corrected *p* < 0.05; Figure 6a-l). Apart from nodal network metrics of left M1, alterations in FC of left M1 were also causally associated with network metrics of other nodes (Supplementary Figure 4). Because nodal DC in left M1 was correlated with most nodal metrics (FDR-corrected *p* < 0.05; Figure 5a), we also examined whether nodal DC in left M1 mediated the effects of striatum SBR on nodal network metrics. Indeed, DC of left M1 mediated the effects of striatum SBR on multiple nodal network metrics (Supplementary Figure 5).

**Figure 6.**
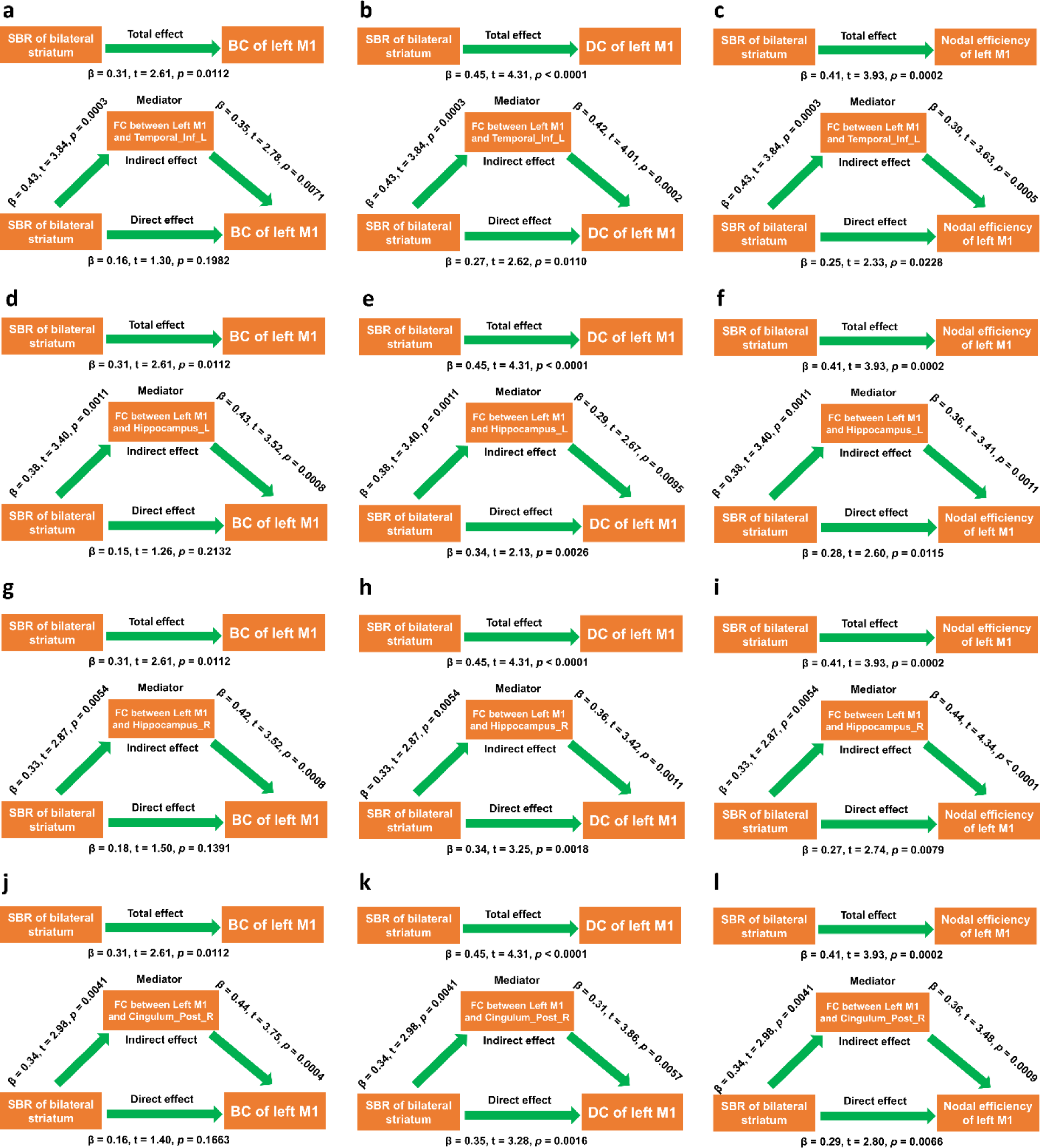
FC of left M1 mediated the effects of striatal dopamine depletion on topological network metrics of left M1. (a-c) FC between left M1 and left inferior temporal gyrus mediated the effects of striatum SBR on BC, DC, and nodal efficiency of left M1. (d-f) FC between left M1 and left hippocampus mediated the effects of striatum SBR on BC, DC, and nodal efficiency of left M1. (g-i) FC between left M1 and right hippocampus mediated the effects of striatum SBR on BC, DC, and nodal efficiency of left M1. (j-l) FC between left M1 and right posterior cingulate cortex mediated the effects of striatum SBR on BC, DC, and nodal efficiency of left M1. Mediation analysis was performed with age, sex, disease duration, and years of education as confounding variables. *p* < 0.05 was considered statistically significant. Abbreviations: BC, Betweenness centrality; DC, Degree centrality; M1, Primary motor cortex; FC, Functional connectivity; SBR, Striatal binding ratio.

### The associations between FC of left M1 and motor symptoms and overall disease burden

The FC between left M1 and left supplementary motor area (SMA) was negatively correlated with UPDRS-III scores (Figure 7a) and mediated the effects of putamen SBR (Figure 7b) and striatum SBR (Figure 7c) on UPDRS-III scores. In addition, the FC between left M1 and left SMA was negatively correlated with total UPDRS scores (Figure 7d) and mediated the effects of putamen SBR (Figure 7e) and striatum SBR (Figure 7f) on total UPDRS scores.

**Figure 7.**
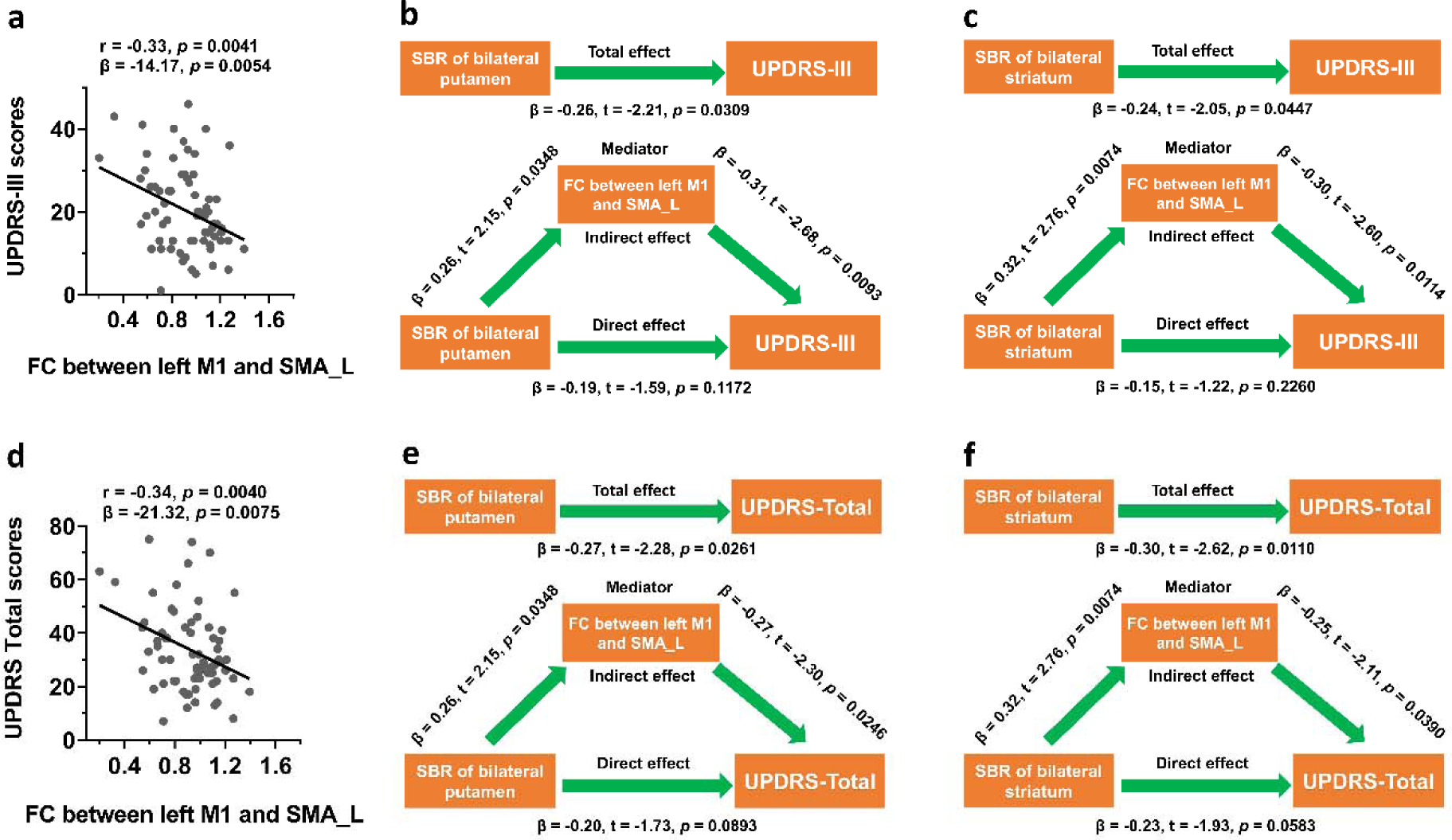
FC of left M1 mediated the effects of striatal dopamine depletion on motor impairment and overall disease burden. (a-c) FC between left M1 and left SMA was negatively correlated with UPDRS-III scores (a) and mediated the effects of bilateral putamen (b) and bilateral striatum (c) on UPDRS-III scores. (d-f) FC between left M1 and left SMA was negatively correlated with total UPDRS scores (d) and mediated the effects of bilateral putamen (e) and bilateral striatum (f) on total UPDRS scores. The correlation analysis was conducted by Pearson correlation method. Mediation analysis was performed with age, sex, disease duration, and years of education as confounding variables. *p* < 0.05 was considered statistically significant. Abbreviations: UPDRS, Unified Parkinson’s Disease Rating Scale; UPDRS-III, Unified Parkinson’s Disease Rating Scale part III; M1, Primary motor cortex; FC, Functional connectivity; SMA, Supplementary motor area; SBR, Striatal binding ratio.

### The associations between FC of left M1 and non-motor symptoms

The FCs between left M1 and right postcentral gyrus and right supramarginal gyrus were positively associated with LNS scores (FDR-corrected *p* < 0.05; Supplementary Figure 6a-b). The FCs between left M1 and left superior temporal gyrus, right superior temporal gyrus, right supramarginal gyrus were positively associated with SDMT scores (FDR-corrected *p* < 0.05; Supplementary Figure 6c-e). The FCs between left M1 and right M1 and right postcentral gyrus were negatively associated with RBDSQ scores (FDR-corrected *p* < 0.05; Supplementary Figure 6f-g). The FCs between left M1 and left middle cingulate gyrus and right middle cingulate gyrus were negatively associated with SCOPA-AUT scores (FDR-corrected *p* < 0.05; Supplementary Figure 6h-i). With mediation analysis, we demonstrated that the FCs of left M1 mediated the effects of striatal dopamine depletion on scores of LNS, SDMT, RBDSQ, and SCOPA-AUT (Figure 8).

**Figure 8.**
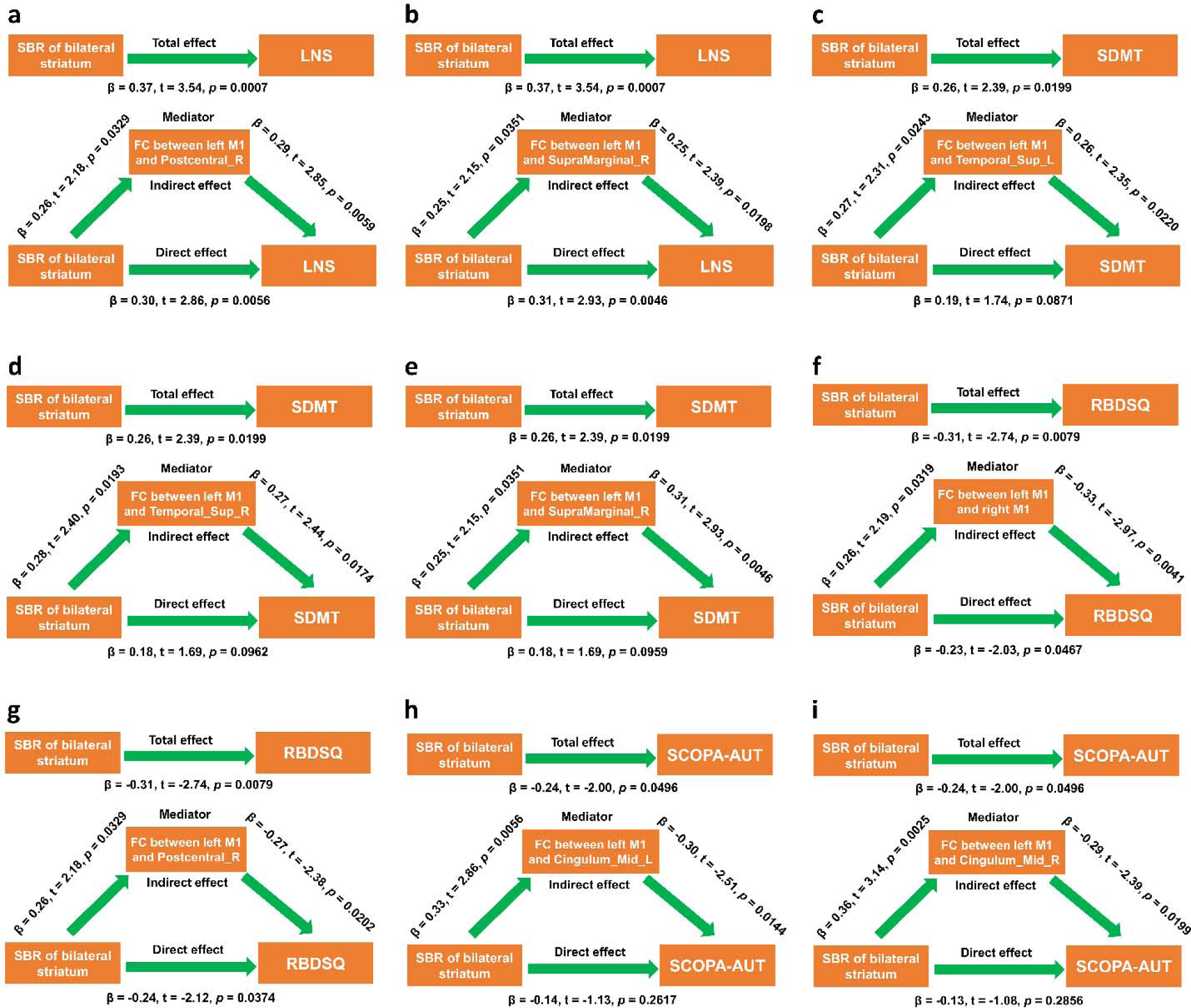
FC of left M1 mediated the effects of striatal dopamine depletion on non-motor assessments. (a-b) FC between left M1 and right postcentral gyrus and right supramarginal gyrus mediated the effects of bilateral striatum on LNS scores. (c-e) FC between left M1 and left superior temporal gyrus, right superior temporal gyrus, right supramarginal gyrus mediated the effects of bilateral striatum on SDMT scores. (f-g) FC between left M1 and right M1 and right postcentral gyrus mediated the effects of bilateral striatum on RBDSQ scores. (h-i) FC between left M1 and left middle cingulate gyrus and right middle cingulate gyrus mediated the effects of bilateral striatum on SCOPA-AUT scores. Mediation analysis was performed with age, sex, disease duration, and years of education as confounding variables. *p* < 0.05 was considered statistically significant. Abbreviations: RBDSQ, REM Sleep Behavior Disorder Screening Questionnaire; SCOPA-AUT, Scale for Outcomes in Parkinson’s Disease-Autonomic; LNS, Letter Number Sequencing test; SDMT, Symbol Digit Modalities Test; FC, Functional connectivity; M1, Primary motor cortex.

### The associations between topological metrics and clinical assessments

For structural network, no significant associations were observed between structural network metrics and clinical assessments of PD patients (all *p* > 0.05 in both Pearson correlation and multivariate regression analysis). For functional network metrics, nodal DC and efficiency of right posterior cingulate cortex were significantly associated with scores of SDMT, RBDSQ, and SCOPA-AUT in PD patients (Supplementary Figure 7). As shown in Supplementary Figure 8, with mediation analysis, we further demonstrated that nodal DC and efficiency of right posterior cingulate cortex mediated the effects of striatal dopamine depletion on scores of SDMT, RBDSQ, and SCOPA-AUT (Supplementary Figure 8).

## Discussion

Striatal dopamine depletion is a key hallmark of PD and contributes to the development of both motor and non-motor symptoms.^5,7,8,10^ The loss of midbrain dopamine neurons is associated with the impairment of initiation, speed, and fluidity of voluntary movement.^50^ It is generally hypothesized that striatal dopamine depletion causes dysfunction of basal ganglia-thalamocortical circuit, which results in the occurrence of motor impairment in PD.^51,52^ Consistent with these findings, we revealed striatum SBR was associated with the severity of motor symptoms, such as tremor, rigidity, and UPDRS-III scores. Indeed, recent studies supported that rigidity was associated with striatum SBR,^7,8^ however, they didn’t observe a statistical correlation between striatum SBR and tremor severity.^7,8^ Therefore, they hypothesized that other neural systems besides the nigrostriatal dopaminergic system may contribute to the generation of resting tremor in PD. Controversially, Fois *et al*. (2021) observed a significant association between parkinsonian resting tremor and contralateral reduction of striatum SBR.^9^ In our study, we revealed a positive association between striatum SBR and tremor severity. Specifically, both SBRs of right caudate (r = 0.20, *p* = 0.0161) and left caudate (r = 0.28, *p* = 0.0009) were positively correlated with tremor scores. The most likely explanation for above discrepancy was that striatum SBR was much higher in PD patients with tremor (tremor dominant) than PD patients without tremor according to previous studies,^53,54^ which naturally resulted in the positive correlation bias between striatum SBR and tremor scores. Future studies were required to decipher how striatum SBR differentially affected motor symptoms of PD and the underlying neural mechanisms.

Excessive daytime sleepiness (EDS) was one of the most prevalent non-motor features in PD.^55–58^ According to a recent meta-analysis, almost 35% of PD patients had EDS and EDS was linked to worse motor and autonomic dysfunction, and higher burden of neuropsychiatric symptoms.^58^ In a recent study, we revealed that PD patients with EDS exhibited lower striatum SBR compared to PD patients without EDS.^59^ In current study, striatum SBR was negatively associated with ESS scores, indicating that striatal dopamine depletion was associated with EDS in PD. Indeed, our result was in agreement with the finding from a previous study showing that lower caudate SBR was correlated with higher EDS scores.^60^ RBD was also one of the most frequent non-motor manifestations in PD and occurred in 42.3% of PD patients.^61,62^ RBD has been considered as a prodromal PD due to its high likelihood to convert into neurodegenerative synucleinopathies, such as PD and dementia with Lewy bodies.^63^ Additionally, RBD in PD was also associated with worse non-motor symptoms, such as cognitive impairment,^64,65^ constipation,^66,67^ hallucination,^66^ and depression.^66^ In current study, striatum SBR was negatively associated with RBDSQ scores, indicating striatal dopamine depletion may contribute to the occurrence of RBD in PD. Consistently, recent studies actually showed that striatum SBR was much lower in PD patients with RBD compared to patients without RBD.^68,69^ Autonomic dysfunction was prevalent in PD patients and was associated with worse motor and non-motor symptoms.^70–72^ In this study, striatum SBR was negatively associated with SCOPA-AUT scores, which was consistent with previous studies showing autonomic dysfunction in PD was associated with striatal dopamine depletion.^18–21^ Cognitive impairment was also one of the most common non-motor features of PD patients ^73^ and associated with disease progression and outcome.^73–75^ Cognitive impairment could occur at the early stage and even prodromal phase of PD.^73,76–78^ According to previous studies, PD patients could exhibit impairments of multiple cognitive domains, including verbal memory,^79–81^ executive function,^82,83^ visuospatial function,^84,85^ episodic memory,^86–88^ and processing speed.^11,89^ In this study, we revealed striatum SBR was positively associated with scores of LNS and SDMT, which indicated that higher striatum SBR was associated with better cognitive function. Consistently, a recent study revealed that dopamine depletion in caudate was related to lower memory performance while dopamine depletion in the anterior putamen was linked to poorer attention performance.^90^ Additionally, striatum SBR was also associated with processing speed,^11^ frontal/executive function,^13,91^ and visuospatial function.^13,91^ Taken together, striatum SBR was significantly associated with a series of non-motor symptoms, including EDS, RBD, autonomic dysfunction, and cognitive impairment.

The aberrations of network topology have been reported by a multitude of neuroimaging studies in PD ^92–97^ and significantly correlated with motor and non-motor symptoms of PD patients.^35,98–101^ In PD, striatal dopamine deficiency is associated with significant alterations of brain functional networks in PD ^30,31^, which can be normalized by dopamine replacement.^32^ Moreover, striatum SBR was also associated with significant changes of structural network in PD.^33^ Therefore, striatum SBR may be also associated with network topology in PD patients. In our study, both nodal network metrics in structural network and functional network were significantly different among different SBR quartile groups. In structural network, PD patients in Q4 group exhibited lower nodal networks in most nodes compared to patients in Q1 group. Among these nodal metrics, nodal Cp of right supramarginal gyrus, nodal efficiency of right superior temporal gyrus, and nodal local efficiency of right rolandic operculum were negatively correlated with SBRs of bilateral striatum. These results indicated that striatum SBR significantly shaped the local topology of structural network in PD. In functional network, patients with higher striatum SBR (Q4 group) showed both lower and higher nodal network metrics compared to patients with lower striatum SBR (Q1 group). More importantly, striatum SBR showed stronger associations with topological metrics in functional network than structural network, indicating striatum SBR substantially shaped functional network topology. Based on these results, we examined whether aberrations of FC in functional network mediated the effects of striatal dopamine depletion on functional network topology. Actually, the global network strengths in functional network were extremely lower in Q1 group than Q4 group, suggesting that striatal dopamine depletion induced tremendous dysfunctions in functional network of PD patients. Importantly, we revealed a subnetwork with left M1 as core node was specifically impaired by striatal dopamine depletion, characterized by substantial decline of FC between left M1 and a number of cortical and subcortical regions. Thus, we hypothesized that the impaired left M1 subnetwork contributed to dysregulations of functional network topology in Q1 group. The evidence for this hypothesis included: (i) FC of left M1 was significantly correlated with altered nodal network metrics; (ii) FC between left M1 and multiple nodes mediated the effects of striatal dopamine depletion on nodal network metrics of left M1; (iii) FC between left M1 and multiple nodes mediated the effects of striatal dopamine depletion on other nodal network metrics; (iv) nodal DC (the number of edges) of left M1 also was significantly correlated with other nodal network metrics and mediated the effects of striatal dopamine depletion on these nodal network metrics. Taken together, we concluded that striatal dopamine depletion induced abnormal functional network topology by impairing left M1 subnetwork. The mechanisms underlying the effects of dopamine on M1 subnetwork were largely unknown. According to previous literature, dopamine has been shown to modulate M1 physiology at the behavioral, network, cellular, and synaptic levels.^102^ Mechanistically, dopamine could control pyramidal neuron activity of M1 via D2 receptors.^103^ However, how and why dopamine enhanced FC of left M1 remain elusive. Future studies are required to decode the molecular mechanisms underlying the effects of dopamine on M1 subnetwork.

In physiological condition, M1 played a key role in generating neural impulses that regulate movements, whereas PD patients presented decreased activity in M1 during movement.^104^ Additionally, the FC of M1 was found to be reduced in both drug naïve PD and PD tested off dopaminergic medication during motor tasks.^105–107^ Specifically, blood oxygenation level-dependent responses of M1 and SMA were significantly reduced in PD patients compared to control individuals.^108^ With disease progression, PD patients exhibited a progressive decrease in regional homogeneity of M1.^109^ In addition, PD patients also displayed reduced fractional amplitude of low-frequency fluctuation in M1 compared to control.^110^ Importantly, M1 is responsive to dopaminergic medication and aberrant FC in M1 can be normalized with the administration of levodopa.^111^ In this study, FC between left M1 and left SMA was negatively associated with UPDRS-III scores and mediated the effects of striatum SBR on UPDRS-III scores, indicating that impaired FC of left M1 was causally associated with motor impairment induced by striatal dopamine depletion. However, there were no significant associations between topological metrics of left M1 and motor symptoms, such as UPDRS-III scores, indicating that the effects of striatum SBR on graphical metrics of M1 were independent of motor phenotypes of PD patients. In addition, we also found no significant associations between other nodal network metrics and motor symptoms, further implying that the nodal network metrics affected by striatum SBR was not related to motor symptoms of PD patients. Taken together, our findings suggested that it was impaired FC of left M1 but not network topology in functional network that contribute to motor symptoms induced by striatal dopamine depletion in PD patients.

The neural mechanisms underlying the effects of striatum SBR on cognitive function of PD patients were largely unknown. In this study, we revealed FC in left M1 was positively associated with LNS and SDMT scores and mediated the effects of striatal dopamine depletion on cognitive impairment, suggesting impaired left M1 subnetwork contributed to cognitive impairment. Moreover, we revealed nodal DC and nodal efficiency of right posterior cingulate cortex were negatively associated with SDMT scores and mediated the effects of striatum SBR on SDMT scores, indicating that enhanced local topology of right posterior cingulate cortex induced by striatal dopamine depletion also contributed to cognitive impairment in PD patients. Posterior cingulate cortex was a key node in default mode network ^112^ and essential for executive, mnemonic and spatial processing functions.^113^ In agreement with our findings, increased FC of posterior cingulate cortex in PD patients with cognitive impairment has been reported.^114^ Based on these results, we hypothesized that increased FC of right posterior cingulate cortex may elevate its DC and nodal efficiency, which contributed to cognitive impairment in PD patients.

The neural correlates underlying the effects of striatum SBR on RBD were still not identified. In this study, we showed FCs in left M1 were negatively associated with RBDSQ scores and mediated the effects of striatal dopamine depletion on RBDSQ scores, suggesting impaired left M1 subnetwork contributed to RBD severity in PD. What’s more, we revealed higher nodal DC and efficiency of right posterior cingulate cortex mediated the effects of striatal dopamine depletion on RBDSQ scores. Although posterior cingulate cortex has been shown to be involved in cognitive impairment of isolated RBD,^115,116^ its role in the occurrence of RBD in PD remain unclear. Future studies were required to decipher the specific mechanisms underlying the negative associations between striatum SBR and RBD in PD patients.

Previous studies reported that striatum SBR was associated with autonomic dysfunction in PD patients,^18–21^ which was consistent with our findings observed in current study. We revealed FC between left M1 and bilateral middle cingulate gyri mediated the effects of striatal dopamine depletion on autonomic dysfunction, suggesting impaired left M1 subnetwork also contributed to autonomic dysfunction in PD patients. In addition, nodal DC and efficiency of right posterior cingulate cortex also mediated the effects of striatum SBR on autonomic dysfunction. In agreement with these findings, white matter density in bilateral cingulate have been revealed to be negatively associated with SCOPA-AUT scores.^117^ Future studies were needed to understand how aberrations cingulate cortices contributed to autonomic dysfunction in PD patients.

## Limitations

In current study, we replicated previous findings that striatum SBR was associated with both motor and non-motor symptoms of PD patients, which was required to be further validated in longitudinal studies. Although it was impaired left M1 subnetwork mediated the effects of striatal dopamine depletion on both functional network topology and motor and non-motor symptoms, the molecular mechanisms underlying these effects were still unclear and deserved to be further explored. In addition, the structural substrates mediating the effects of striatal dopamine depletion on structural network topology also remain to be identified in future studies. Furthermore, although nodal DC and efficiency of right posterior cingulate cortex mediated the associations between striatum SBR and non-motor symptoms, how right posterior cingulate cortex contributed the occurrence of non-motor symptoms in PD was poorly understood. More importantly, because left M1 subnetwork contributed to abnormal network topology and more severe disease manifestations, future studies were required to confirm whether neuromodulation of left M1 had therapeutic value in PD patients.

## Contributors

Zhichun Chen, Conceptualization, Formal analysis, Visualization, Methodology, Writing - original draft, Writing - review and editing; Guanglu Li, Data curation, Formal analysis, Visualization; Liche Zhou, Data curation, Formal analysis, Investigation; Lina Zhang, Formal analysis, Investigation, Methodology; Jun Liu, Conceptualization, Supervision, Funding acquisition, Writing - original draft, Project administration, Writing-review and editing. All authors read and approved the final version of the manuscript.

## Data Availability

All the raw data used in the preparation of this Article were downloaded from PPMI database (www.ppmi-info.org/data).All data produced in the present study are available upon reasonable request to the authors.

https://www.ppmi-info.org/data

## Acknowledgments

This work was supported by grants from National Natural Science Foundation of China (Grant No. 81873778, 82071415) and National Research Center for Translational Medicine at Shanghai, Ruijin Hospital, Shanghai Jiao Tong University School of Medicine (Grant No. NRCTM(SH)-2021-03). Data used in the preparation of this article were obtained from the Parkinson’s Progression Markers Initiative (PPMI) database (www.ppmiinfo.org/data). We thank the share of PPMI data by all the PPMI study investigators. PPMI – a public-private partnership – is funded by the Michael J. Fox Foundation for Parkinson’s Research and funding partners, which can be found at www.ppmiinfo.org/fundingpartners.

## Supplementary materials

Additional supporting information may be found online in the supplementary materials section at the end of the article.

## References

1 Armstrong MJ, Okun MS. Diagnosis and Treatment of Parkinson Disease: A Review. JAMA. 2020; 323(6): 548–60.

2 Bloem BR, Okun MS, Klein C. Parkinson’s disease. Lancet. 2021; 397(10291): 2284–303.

3 Michel PP, Hirsch EC, Hunot S. Understanding Dopaminergic Cell Death Pathways in Parkinson Disease. Neuron. 2016; 90(4): 675–91.

4 Cramb KML, Beccano-Kelly D, Cragg SJ, Wade-Martins R. Impaired dopamine release in Parkinson’s disease. Brain. 2023; 146(8): 3117–32.

5 Heng N, Malek N, Lawton MA, et al. Striatal Dopamine Loss in Early Parkinson’s Disease: Systematic Review and Novel Analysis of Dopamine Transporter Imaging. Mov Disord Clin Pract. 2023; 10(4): 539–46.

6 Simuni T, Uribe L, Cho HR, et al. Clinical and dopamine transporter imaging characteristics of non-manifest LRRK2 and GBA mutation carriers in the Parkinson’s Progression Markers Initiative (PPMI): a cross-sectional study. Lancet Neurol. 2020; 19(1): 71–80.

7 Makinen E, Joutsa J, Jaakkola E, et al. Individual parkinsonian motor signs and striatal dopamine transporter deficiency: a study with [I-123]FP-CIT SPECT. J Neurol. 2019; 266(4): 826–34.

8 Moccia M, Pappata S, Picillo M, et al. Dopamine transporter availability in motor subtypes of de novo drug-naive Parkinson’s disease. J Neurol. 2014; 261(11): 2112–8.

9 Fois AF, Chang FC, Barnett R, et al. Rest tremor correlates with reduced contralateral striatal dopamine transporter binding in Parkinson’s disease. Parkinsonism Relat Disord. 2021; 85: 102–8.

10 Liu R, Umbach DM, Troster AI, Huang X, Chen H. Non-motor symptoms and striatal dopamine transporter binding in early Parkinson’s disease. Parkinsonism Relat Disord. 2020; 72: 23–30.

11 Vriend C, van Balkom TD, van Druningen C, et al. Processing speed is related to striatal dopamine transporter availability in Parkinson’s disease. Neuroimage Clin. 2020; 26: 102257.

12 Fiorenzato E, Antonini A, Bisiacchi P, Weis L, Biundo R. Asymmetric Dopamine Transporter Loss Affects Cognitive and Motor Progression in Parkinson’s Disease. Mov Disord. 2021; 36(10): 2303–13.

13 Chung SJ, Yoo HS, Oh JS, et al. Effect of striatal dopamine depletion on cognition in de novo Parkinson’s disease. Parkinsonism Relat Disord. 2018; 51: 43–8.

14 Costello H, Yamamori Y, Reeves S, Schrag AE, Howard R, Roiser JP. Longitudinal decline in striatal dopamine transporter binding in Parkinson’s disease: associations with apathy and anhedonia. J Neurol Neurosurg Psychiatry. 2023.

15 Vriend C, Raijmakers P, Veltman DJ, et al. Depressive symptoms in Parkinson’s disease are related to reduced [123I]FP-CIT binding in the caudate nucleus. J Neurol Neurosurg Psychiatry. 2014; 85(2): 159–64.

16 Yoo SW, Oh YS, Hwang EJ, et al. “Depressed” caudate and ventral striatum dopamine transporter availability in de novo Depressed Parkinson’s disease. Neurobiol Dis. 2019; 132: 104563.

17 Vriend C, Nordbeck AH, Booij J, et al. Reduced dopamine transporter binding predates impulse control disorders in Parkinson’s disease. Mov Disord. 2014; 29(7): 904–11.

18 Contaldi E, Magistrelli L, Gallo S, Comi C. Striatal dopamine transporter imaging in Parkinson’s disease drug-naive patients: focus on sexual dysfunction. Neurol Sci. 2022; 43(8): 4769–76.

19 Mito Y, Yabe I, Yaguchi H, Takei T, Terae S, Tajima Y. Relation of overactive bladder with motor symptoms and dopamine transporter imaging in drug-naive Parkinson’s disease. Parkinsonism Relat Disord. 2018; 50: 37–41.

20 Yoshii F, Ryo M, Baba Y, Koide T, Hashimoto J. Combined use of dopamine transporter imaging (DAT-SPECT) and (123)I-metaiodobenzylguanidine (MIBG) myocardial scintigraphy for diagnosing Parkinson’s disease. J Neurol Sci. 2017; 375: 80–5.

21 Murtomaki K, Mertsalmi T, Jaakkola E, et al. Gastrointestinal Symptoms and Dopamine Transporter Asymmetry in Early Parkinson’s Disease. Mov Disord. 2022; 37(6): 1284–9.

22 Nabizadeh F, Sodeifian F, Pirahesh K. Olfactory dysfunction and striatal dopamine transporter binding in motor subtypes of Parkinson’s disease. Neurol Sci. 2022; 43(8): 4745–52.

23 Noudoost B, Moore T. Control of visual cortical signals by prefrontal dopamine. Nature. 2011; 474(7351): 372–5.

24 Vander Weele CM, Siciliano CA, Matthews GA, et al. Dopamine enhances signal-to-noise ratio in cortical-brainstem encoding of aversive stimuli. Nature. 2018; 563(7731): 397–401.

25 Zaldivar D, Rauch A, Whittingstall K, Logothetis NK, Goense J. Dopamine-induced dissociation of BOLD and neural activity in macaque visual cortex. Curr Biol. 2014; 24(23): 2805–11.

26 Calabro FJ, Montez DF, Larsen B, et al. Striatal dopamine supports reward expectation and learning: A simultaneous PET/fMRI study. Neuroimage. 2023; 267: 119831.

27 Kahnt T, Tobler PN. Dopamine Modulates the Functional Organization of the Orbitofrontal Cortex. J Neurosci. 2017; 37(6): 1493–504.

28 Dang LC, Donde A, Madison C, O’Neil JP, Jagust WJ. Striatal dopamine influences the default mode network to affect shifting between object features. J Cogn Neurosci. 2012; 24(9): 1960–70.

29 Nagano-Saito A, Leyton M, Monchi O, Goldberg YK, He Y, Dagher A. Dopamine depletion impairs frontostriatal functional connectivity during a set-shifting task. J Neurosci. 2008; 28(14): 3697–706.

30 Shine JM, Bell PT, Matar E, et al. Dopamine depletion alters macroscopic network dynamics in Parkinson’s disease. Brain. 2019; 142(4): 1024–34.

31 Dirkx MF, den Ouden HE, Aarts E, et al. Dopamine controls Parkinson’s tremor by inhibiting the cerebellar thalamus. Brain. 2017; 140(3): 721–34.

32 Wu C, Wu H, Zhou C, et al. Normalization effect of dopamine replacement therapy on brain functional connectome in Parkinson’s disease. Hum Brain Mapp. 2023; 44(9): 3845–58.

33 Chung SJ, Kim YJ, Kim YJ, et al. Association Between White Matter Networks and the Pattern of Striatal Dopamine Depletion in Patients With Parkinson Disease. Neurology. 2022.

34 Choi H, Cheon GJ, Kim HJ, et al. Gray matter correlates of dopaminergic degeneration in Parkinson’s disease: A hybrid PET/MR study using (18) F-FP-CIT. Hum Brain Mapp. 2016; 37(5): 1710–21.

35 Chen Z, Wu B, Li G, Zhou L, Zhang L, Liu J. Age and sex differentially shape brain networks in Parkinson’s disease. CNS Neurosci Ther. 2023.

36 Zhang L, Yang T, Chen Y, et al. Cognitive Deficit and Aberrant Intrinsic Brain Functional Network in Early-Stage Drug-Naive Parkinson’s Disease. Front Neurosci. 2022; 16: 725766.

37 Ehgoetz Martens KA, Hall JM, Georgiades MJ, et al. The functional network signature of heterogeneity in freezing of gait. Brain. 2018; 141(4): 1145–60.

38 Chen Z, Wu B, Li G, Zhou L, Zhang L, Liu J. Parkinson’s disease-associated genetic variants synergistically shape brain networks. medRxiv. 2023: 2022.12.25.22283938.

39 Chen Z, Wu B, Li G, Zhou L, Zhang L, Liu J. BCKDK rs14235 A allele is associated with milder motor impairment and altered network topology in Parkinson’s disease. medRxiv. 2023: 2023.07.20.23292985.

40 Parkinson Progression Marker I. The Parkinson Progression Marker Initiative (PPMI). Prog Neurobiol. 2011; 95(4): 629–35.

41 Marek K, Chowdhury S, Siderowf A, et al. The Parkinson’s progression markers initiative (PPMI) - establishing a PD biomarker cohort. Ann Clin Transl Neurol. 2018; 5(12): 1460–77.

42 Hol L, Van Oosten P, Nijbroek S, et al. The effect of age on ventilation management and clinical outcomes in critically ill COVID-19 patients--insights from the PRoVENT-COVID study. Aging (Albany NY). 2022; 14(3): 1087–109.

43 Oliveira JCS, Santos AMD, Aguiar MF, et al. Characteristics of Older Patients with Takayasu’s Arteritis: A Two-Center, Cross-Sectional, Retrospective Cohort Study. Arq Bras Cardiol. 2023; 120(1): e20220463.

44 Yue X, Li Z, Li Y, et al. Altered Static and Dynamic Functional Network Connectivity in Post-stroke Cognitive Impairment. Neurosci Lett. 2023: 137097.

45 Lesourd M, Reynaud E, Navarro J, et al. Involvement of the posterior tool processing network during explicit retrieval of action tool and semantic tool knowledge: an fMRI study. Cereb Cortex. 2023.

46 Chaddock-Heyman L, Weng TB, Kienzler C, et al. Scholastic performance and functional connectivity of brain networks in children. PLoS One. 2018; 13(1): e0190073.

47 Wu GR, Stramaglia S, Chen H, Liao W, Marinazzo D. Mapping the voxel-wise effective connectome in resting state FMRI. PLoS One. 2013; 8(9): e73670.

48 Rubinov M, Sporns O. Complex network measures of brain connectivity: uses and interpretations. Neuroimage. 2010; 52(3): 1059–69.

49 Wang JH, Zuo XN, Gohel S, Milham MP, Biswal BB, He Y. Graph theoretical analysis of functional brain networks: test-retest evaluation on short- and long-term resting-state functional MRI data. PLoS One. 2011; 6(7): e21976.

50 Panigrahi B, Martin KA, Li Y, et al. Dopamine Is Required for the Neural Representation and Control of Movement Vigor. Cell. 2015; 162(6): 1418–30.

51 Obeso JA, Marin C, Rodriguez-Oroz C, et al. The basal ganglia in Parkinson’s disease: current concepts and unexplained observations. Ann Neurol. 2008; 64 Suppl 2: S30–46.

52 McGregor MM, Nelson AB. Circuit Mechanisms of Parkinson’s Disease. Neuron. 2019; 101(6): 1042–56.

53 Lee JY, Lao-Kaim NP, Pasquini J, Deuschl G, Pavese N, Piccini P. Pallidal dopaminergic denervation and rest tremor in early Parkinson’s disease: PPMI cohort analysis. Parkinsonism Relat Disord. 2018; 51: 101–4.

54 Kaasinen V, Kinos M, Joutsa J, Seppanen M, Noponen T. Differences in striatal dopamine transporter density between tremor dominant and non-tremor Parkinson’s disease. Eur J Nucl Med Mol Imaging. 2014; 41(10): 1931–7.

55 Tholfsen LK, Larsen JP, Schulz J, Tysnes OB, Gjerstad MD. Development of excessive daytime sleepiness in early Parkinson disease. Neurology. 2015; 85(2): 162–8.

56 Feng F, Cai Y, Hou Y, Ou R, Jiang Z, Shang H. Excessive daytime sleepiness in Parkinson’s disease: A systematic review and meta-analysis. Parkinsonism Relat Disord. 2021; 85: 133–40.

57 Amara AW, Chahine LM, Caspell-Garcia C, et al. Longitudinal assessment of excessive daytime sleepiness in early Parkinson’s disease. J Neurol Neurosurg Psychiatry. 2017; 88(8): 653–62.

58 Maggi G, Vitale C, Cerciello F, Santangelo G. Sleep and wakefulness disturbances in Parkinson’s disease: A meta-analysis on prevalence and clinical aspects of REM sleep behavior disorder, excessive daytime sleepiness and insomnia. Sleep Med Rev. 2023; 68: 101759.

59 Chen Z, Wu B, Li G, Zhou L, Zhang L, Liu J. BIN3 rs2280104 T allele is associated with excessive daytime sleepiness and altered network topology in Parkinson’s disease. medRxiv. 2023: 2023.07.17.23292760.

60 Yousaf T, Pagano G, Niccolini F, Politis M. Excessive daytime sleepiness may be associated with caudate denervation in Parkinson disease. J Neurol Sci. 2018; 387: 220–7.

61 Adler CH, Hentz JG, Shill HA, et al. Probable RBD is increased in Parkinson’s disease but not in essential tremor or restless legs syndrome. Parkinsonism Relat Disord. 2011; 17(6): 456–8.

62 Zhang X, Sun X, Wang J, Tang L, Xie A. Prevalence of rapid eye movement sleep behavior disorder (RBD) in Parkinson’s disease: a meta and meta-regression analysis. Neurol Sci. 2017; 38(1): 163–70.

63 Fereshtehnejad SM, Montplaisir JY, Pelletier A, Gagnon JF, Berg D, Postuma RB. Validation of the MDS research criteria for prodromal Parkinson’s disease: Longitudinal assessment in a REM sleep behavior disorder (RBD) cohort. Mov Disord. 2017; 32(6): 865–73.

64 Oltra J, Uribe C, Segura B, et al. Brain atrophy pattern in de novo Parkinson’s disease with probable RBD associated with cognitive impairment. NPJ Parkinsons Dis. 2022; 8(1): 60.

65 Yan YY, Lei K, Li YY, Liu XF, Chang Y. The correlation between possible RBD and cognitive function in Parkinson’s disease patients in China. Ann Clin Transl Neurol. 2019; 6(5): 848–53.

66 Xie D, Shen Q, Zhou J, Xu Y. Non-motor symptoms are associated with REM sleep behavior disorder in Parkinson’s disease: a systematic review and meta-analysis. Neurol Sci. 2021; 42(1): 47–60.

67 Chen Y, Xu Q, Wu L, et al. REM sleep behavior disorder correlates with constipation in de novo Chinese Parkinson’s disease patients. Neurol Sci. 2023; 44(1): 191–7.

68 Kwak IH, Lee YK, Ma HI, et al. Striatal Subregion Analysis Associated with REM Sleep Behavior Disorder in Parkinson’s Disease. J Integr Neurosci. 2023; 22(1): 18.

69 Xu Q, Jiang C, Ge J, et al. The impact of probable rapid eye movement sleep behavior disorder on Parkinson’s disease: A dual-tracer PET imaging study. Parkinsonism Relat Disord. 2022; 95: 47–53.

70 Zhou Z, Zhou X, Zhou X, et al. Characteristics of Autonomic Dysfunction in Parkinson’s Disease: A Large Chinese Multicenter Cohort Study. Front Aging Neurosci. 2021; 13: 761044.

71 Chen Z, Li G, Liu J. Autonomic dysfunction in Parkinson’s disease: Implications for pathophysiology, diagnosis, and treatment. Neurobiol Dis. 2020; 134: 104700.

72 De Pablo-Fernandez E, Tur C, Revesz T, Lees AJ, Holton JL, Warner TT. Association of Autonomic Dysfunction With Disease Progression and Survival in Parkinson Disease. JAMA Neurol. 2017; 74(8): 970–6.

73 Aarsland D, Batzu L, Halliday GM, et al. Parkinson disease-associated cognitive impairment. Nat Rev Dis Primers. 2021; 7(1): 47.

74 Gan Y, Xie H, Qin G, et al. Association between Cognitive Impairment and Freezing of Gait in Patients with Parkinson’s Disease. J Clin Med. 2023; 12(8).

75 Mak MK, Wong A, Pang MY. Impaired executive function can predict recurrent falls in Parkinson’s disease. Arch Phys Med Rehabil. 2014; 95(12): 2390–5.

76 Pan C, Li Y, Ren J, et al. Characterizing mild cognitive impairment in prodromal Parkinson’s disease: A community-based study in China. CNS Neurosci Ther. 2022; 28(2): 259–68.

77 Guner D, Tiftikcioglu BI, Tuncay N, Zorlu Y. Contribution of Quantitative EEG to the Diagnosis of Early Cognitive Impairment in Patients With Idiopathic Parkinson’s Disease. Clin EEG Neurosci. 2017; 48(5): 348–54.

78 Markaki I, Bergstrom S, Tsitsi P, et al. Cerebrospinal Fluid Levels of Kininogen-1 Indicate Early Cognitive Impairment in Parkinson’s Disease. Mov Disord. 2020; 35(11): 2101–6.

79 Chen Z, Wu B, Li G, Zhou L, Zhang L, Liu J. MAPT rs17649553 T allele is associated with better verbal memory and higher small-world properties in Parkinson’s disease. Neurobiol Aging. 2023; 129: 219–31.

80 Hanoglu L, Ercan FB, Mantar N, et al. Accelerated forgetting and verbal memory consolidation process in idiopathic nondement Parkinson’s disease. J Clin Neurosci. 2019; 70: 208–13.

81 Lucas-Jimenez O, Diez-Cirarda M, Ojedaa N, Pena J, Cabrera-Zubizarreta A, Ibarretxe-Bilbao N. Verbal Memory in Parkinson’s Disease: A Combined DTI and fMRI Study. J Parkinsons Dis. 2015; 5(4): 793–804.

82 Cameron IG, Watanabe M, Pari G, Munoz DP. Executive impairment in Parkinson’s disease: response automaticity and task switching. Neuropsychologia. 2010; 48(7): 1948–57.

83 Boon LI, Hepp DH, Douw L, et al. Functional connectivity between resting-state networks reflects decline in executive function in Parkinson’s disease: A longitudinal fMRI study. Neuroimage Clin. 2020; 28: 102468.

84 Kawashima S, Shimizu Y, Ueki Y, Matsukawa N. Impairment of the visuospatial working memory in the patients with Parkinson’s Disease: an fMRI study. BMC Neurol. 2021; 21(1): 335.

85 Artusi CA, Montanaro E, Erro R, et al. Visuospatial Deficits Are Associated with Pisa Syndrome and not Camptocormia in Parkinson’s Disease. Mov Disord Clin Pract. 2023; 10(1): 64–73.

86 Dunet V, Fartaria MJ, Deverdun J, et al. Episodic memory decline in Parkinson’ s disease: relation with white matter hyperintense lesions and influence of quantification method. Brain Imaging Behav. 2019; 13(3): 810–8.

87 La C, Linortner P, Bernstein JD, et al. Hippocampal CA1 subfield predicts episodic memory impairment in Parkinson’s disease. Neuroimage Clin. 2019; 23: 101824.

88 Siquier A, Andres P. Episodic Memory Impairment in Parkinson’s Disease: Disentangling the Role of Encoding and Retrieval. J Int Neuropsychol Soc. 2021; 27(3): 261–9.

89 Cholerton BA, Poston KL, Yang L, et al. Semantic fluency and processing speed are reduced in non-cognitively impaired participants with Parkinson’s disease. J Clin Exp Neuropsychol. 2021; 43(5): 469–80.

90 Fornari LHT, da Silva Junior N, Muratt Carpenedo C, Hilbig A, Rieder CRM. Striatal dopamine correlates to memory and attention in Parkinson’s disease. Am J Nucl Med Mol Imaging. 2021; 11(1): 10–9.

91 Kim H, Oh M, Oh JS, et al. Association of striatal dopaminergic neuronal integrity with cognitive dysfunction and cerebral cortical metabolism in Parkinson’s disease with mild cognitive impairment. Nucl Med Commun. 2019; 40(12): 1216–23.

92 Luo CY, Guo XY, Song W, et al. Functional connectome assessed using graph theory in drug-naive Parkinson’s disease. J Neurol. 2015; 262(6): 1557–67.

93 Zhang D, Wang J, Liu X, Chen J, Liu B. Aberrant Brain Network Efficiency in Parkinson’s Disease Patients with Tremor: A Multi-Modality Study. Front Aging Neurosci. 2015; 7: 169.

94 Guo M, Ren Y, Yu H, et al. Alterations in Degree Centrality and Functional Connectivity in Parkinson’s Disease Patients With Freezing of Gait: A Resting-State Functional Magnetic Resonance Imaging Study. Front Neurosci. 2020; 14: 582079.

95 Ji GJ, Ren C, Li Y, et al. Regional and network properties of white matter function in Parkinson’s disease. Hum Brain Mapp. 2019; 40(4): 1253–63.

96 Fang J, Chen H, Cao Z, et al. Impaired brain network architecture in newly diagnosed Parkinson’s disease based on graph theoretical analysis. Neurosci Lett. 2017; 657: 151–8.

97 Wei L, Zhang J, Long Z, et al. Reduced topological efficiency in cortical-basal Ganglia motor network of Parkinson’s disease: a resting state fMRI study. PLoS One. 2014; 9(10): e108124.

98 Hou Y, Wei Q, Ou R, et al. Impaired topographic organization in cognitively unimpaired drug-naive patients with rigidity-dominant Parkinson’s disease. Parkinsonism Relat Disord. 2018; 56: 52–7.

99 Rucco R, Lardone A, Liparoti M, et al. Brain Networks and Cognitive Impairment in Parkinson’s Disease. Brain Connect. 2022; 12(5): 465–75.

100 Shang S, Zhu S, Wu J, et al. Topological disruption of high-order functional networks in cognitively preserved Parkinson’s disease. CNS Neurosci Ther. 2023; 29(2): 566–76.

101 Suo X, Lei D, Li N, et al. Functional Brain Connectome and Its Relation to Hoehn and Yahr Stage in Parkinson Disease. Radiology. 2017; 285(3): 904–13.

102 Cousineau J, Plateau V, Baufreton J, Le Bon-Jego M. Dopaminergic modulation of primary motor cortex: From cellular and synaptic mechanisms underlying motor learning to cognitive symptoms in Parkinson’s disease. Neurobiol Dis. 2022; 167: 105674.

103 Vitrac C, Peron S, Frappe I, et al. Dopamine control of pyramidal neuron activity in the primary motor cortex via D2 receptors. Front Neural Circuits. 2014; 8: 13.

104 Catalan MJ, Ishii K, Honda M, Samii A, Hallett M. A PET study of sequential finger movements of varying length in patients with Parkinson’s disease. Brain. 1999; 122 (Pt 3): 483–95.

105 Prodoehl J, Spraker M, Corcos D, Comella C, Vaillancourt D. Blood oxygenation level-dependent activation in basal ganglia nuclei relates to specific symptoms in de novo Parkinson’s disease. Mov Disord. 2010; 25(13): 2035–43.

106 Spraker MB, Prodoehl J, Corcos DM, Comella CL, Vaillancourt DE. Basal ganglia hypoactivity during grip force in drug naive Parkinson’s disease. Hum Brain Mapp. 2010; 31(12): 1928–41.

107 Mohl B, Berman BD, Shelton E, Tanabe J. Levodopa response differs in Parkinson’s motor subtypes: A task-based effective connectivity study. J Comp Neurol. 2017; 525(9): 2192–201.

108 Buhmann C, Glauche V, Sturenburg HJ, Oechsner M, Weiller C, Buchel C. Pharmacologically modulated fMRI--cortical responsiveness to levodopa in drug-naive hemiparkinsonian patients. Brain. 2003; 126(Pt 2): 451–61.

109 Zeng Q, Guan X, Law Yan Lun JCF, et al. Longitudinal Alterations of Local Spontaneous Brain Activity in Parkinson’s Disease. Neurosci Bull. 2017; 33(5): 501–9.

110 Hu XF, Zhang JQ, Jiang XM, et al. Amplitude of low-frequency oscillations in Parkinson’s disease: a 2-year longitudinal resting-state functional magnetic resonance imaging study. Chin Med J (Engl). 2015; 128(5): 593–601.

111 Shen Y, Hu J, Chen Y, et al. Levodopa Changes Functional Connectivity Patterns in Subregions of the Primary Motor Cortex in Patients With Parkinson’s Disease. Front Neurosci. 2020; 14: 647.

112 Leech R, Kamourieh S, Beckmann CF, Sharp DJ. Fractionating the default mode network: distinct contributions of the ventral and dorsal posterior cingulate cortex to cognitive control. J Neurosci. 2011; 31(9): 3217–24.

113 Foster BL, Koslov SR, Aponik-Gremillion L, Monko ME, Hayden BY, Heilbronner SR. A tripartite view of the posterior cingulate cortex. Nat Rev Neurosci. 2023; 24(3): 173–89.

114 Zhan ZW, Lin LZ, Yu EH, et al. Abnormal resting-state functional connectivity in posterior cingulate cortex of Parkinson’s disease with mild cognitive impairment and dementia. CNS Neurosci Ther. 2018; 24(10): 897–905.

115 Remillard-Pelchat D, Rahayel S, Gaubert M, et al. Comprehensive Analysis of Brain Volume in REM Sleep Behavior Disorder with Mild Cognitive Impairment. J Parkinsons Dis. 2022; 12(1): 229–41.

116 Vacca M, Assogna F, Pellicano C, et al. Neuropsychiatric, neuropsychological, and neuroimaging features in isolated REM sleep behavior disorder: The importance of MCI. Sleep Med. 2022; 100: 230–7.

117 Ashraf-Ganjouei A, Majd A, Javinani A, Aarabi MH. Autonomic dysfunction and white matter microstructural changes in drug-naive patients with Parkinson’s disease. PeerJ. 2018; 6: e5539.

