## Supplementary materials for "Striatal dopamine depletion drives disease progression and network topology aberrations specifically by impairing left M1 network"

| Clinical variable | Q1 group  (n = 35) | Q2-3 group  (n = 70) | Q4 group  (n = 36) | Q1 *vs* Q2-3 | Q1 *vs* Q4 | Q2-3 *vs* Q4 |
| --- | --- | --- | --- | --- | --- | --- |
| Age, years | 62.22 ± 7.59 | 62.62 ± 9.41 | 57.17 ± 9.22 | *p* > 0.05 | ***p* < 0.05** | ***p* < 0.01** |
| Sex (Male/Female) | 27/8 | 44/26 | 18/18 | *p* > 0.05 | ***p* < 0.05** | *p* > 0.05 |
| Education, years | 15.60 ± 2.49 | 14.74 ± 2.93 | 16.42 ± 3.02 | *p* > 0.05 | *p* > 0.05 | ***p* < 0.05** |
| Disease duration, years | 1.78 ± 1.29 | 2.21 ± 2.57 | 1.88 ± 1.93 | *p* > 0.05 | *p* > 0.05 | *p* > 0.05 |
| SBR of Caudate_R | 1.15 ± 0.26 | 1.80 ± 0.30 | 2.42 ± 0.32 | ***p* < 0.0001** | ***p* < 0.0001** | ***p* < 0.0001** |
| SBR of Caudate_L | 1.11 ± 0.33 | 1.79 ± 0.34 | 2.42 ± 0.42 | ***p* < 0.0001** | ***p* < 0.0001** | ***p* < 0.0001** |
| SBR of Putamen_R | 0.49 ± 0.15 | 0.70 ± 0.19 | 1.06 ± 0.40 | ***p* < 0.001** | ***p* < 0.0001** | ***p* < 0.0001** |
| SBR of Putamen_L | 0.46 ± 0.15 | 0.69 ± 0.19 | 1.08 ± 0.38 | ***p* < 0.0001** | ***p* < 0.0001** | ***p* < 0.0001** |
| SBR of Striatum_R | 1.64 ± 0.37 | 2.50 ± 0.44 | 3.48 ± 0.64 | ***p* < 0.0001** | ***p* < 0.0001** | ***p* < 0.0001** |
| SBR of Striatum_L | 1.56 ± 0.42 | 2.48 ± 0.48 | 3.50 ± 0.73 | ***p* < 0.0001** | ***p* < 0.0001** | ***p* < 0.0001** |
| SBR of Bilateral Caudate | 1.13 ± 0.24 | 1.79 ± 0.24 | 2.42 ± 0.29 | ***p* < 0.0001** | ***p* < 0.0001** | ***p* < 0.0001** |
| SBR of Bilateral Putamen | 0.47 ± 0.11 | 0.70 ± 0.13 | 1.07 ± 0.29 | ***p* < 0.0001** | ***p* < 0.0001** | ***p* < 0.0001** |
| SBR of Bilateral Striatum | 0.80 ± 0.15 | 1.24 ± 0.16 | 1.74 ± 0.25 | ***p* < 0.0001** | ***p* < 0.0001** | ***p* < 0.0001** |

**Supplementary Materials**

***Table S1:* The demographic data and SBRs for each quartile group**

The data were shown as the mean ± standard deviation (SD). One-way ANOVA followed by Tukey’s post hoc test (Q1 group *vs* Q2-3 group *vs* Q4 group) were utilized to compare continuous variables. χ^2^ test was used to compare categorical variable (Sex). *p* < 0.05 was deemed statistically significant. The bold values indicate statistical significance. Abbreviations: SBR, striatal binding ratio.

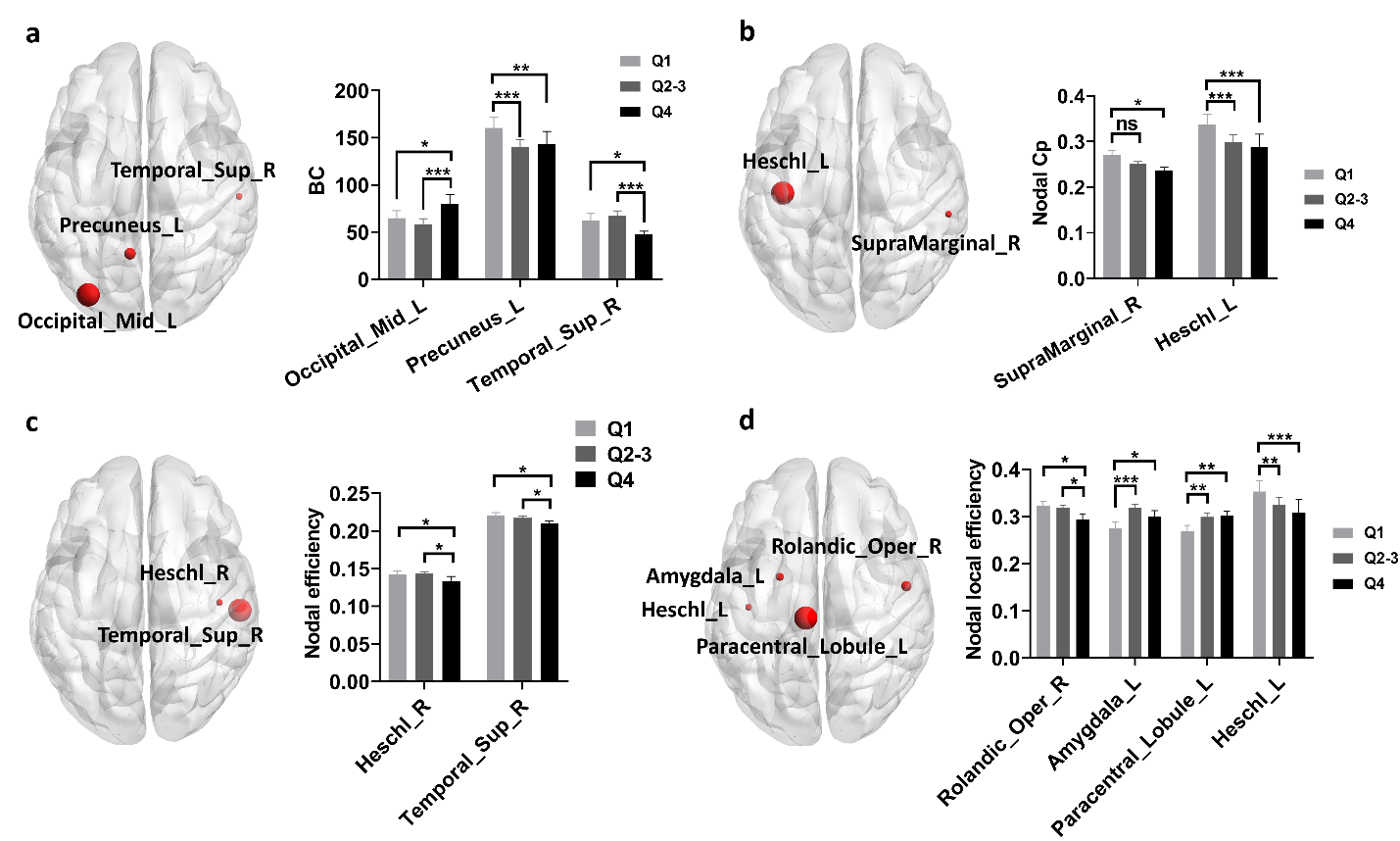

**Supplementary Figure 1. Group differences of nodal network metrics in structural network.** (a-d) Group differences of nodal BC (a), nodal Cp (b), nodal efficiency (c), and nodal local efficiency (d) in structural network among different quartile groups. Two-way ANOVA test with FDR correction was performed. *p* < 0.05 after FDR correction was considered to have statistical significance. **p* < 0.05, ** *p* < 0.01, ****p* < 0.001. Abbreviations: BC, Betweenness centrality; Cp, Clustering coefficient.

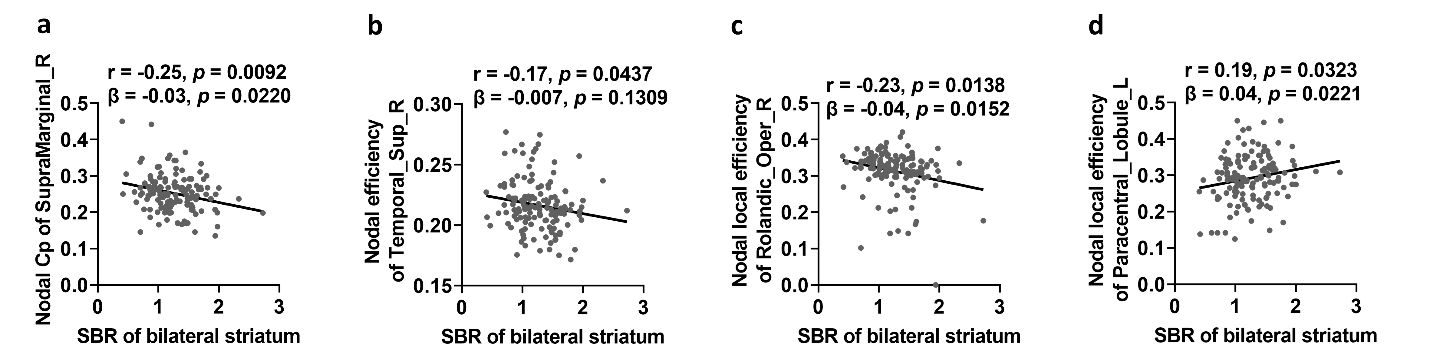

**Supplementary Figure 2. Associations between striatum SBR and nodal network metrics in structural network.** (a-d) Striatum SBR was negatively associated with nodal Cp of right supramarginal gyrus, nodal efficiency of right superior temporal gyrus, nodal local efficiency of right rolandic operculum and positively associated with nodal local efficiency of left paracentral lobule (FDR-corrected *p* < 0.05 in both Pearson correlation analysis and multivariate regression analysis). The association analysis between graphical network metrics and striatum SBR was conducted by Pearson correlation method and multivariate regression analysis with age, sex, disease duration, and years of education as covariates. FDR-corrected *p* < 0.05 was considered statistically significant. Abbreviations: Cp, Clustering coefficient; SBR, Striatal binding ratio.

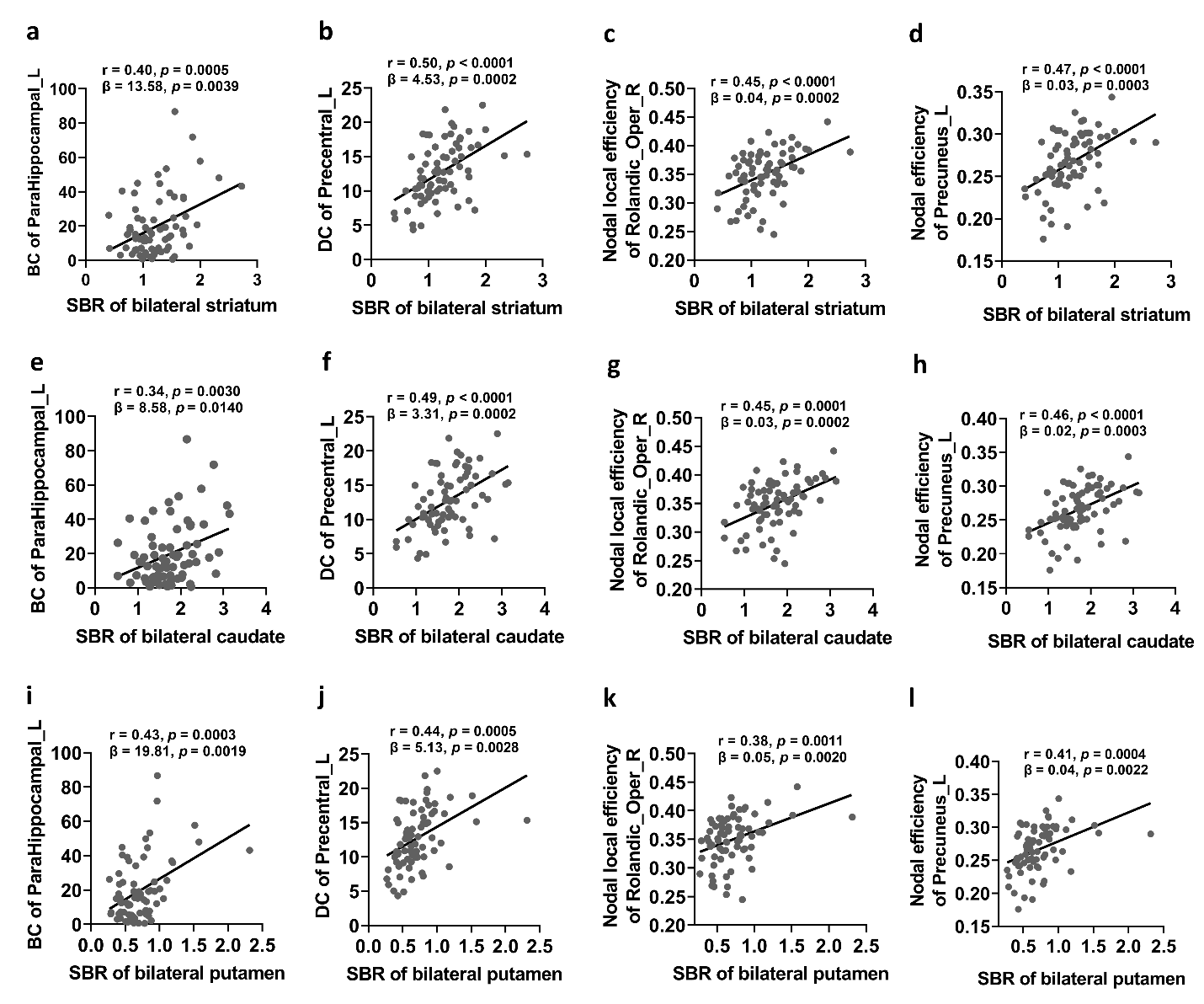

**Supplementary Figure 3. Associations between striatum SBR and nodal network metrics in functional network.** (a-d) Striatum SBR was positively associated with BC of left parahippocampal gyrus, DC of left M1, nodal local efficiency of right rolandic operculum, and nodal efficiency of left M1 (FDR-corrected *p* < 0.05 in both Pearson correlation analysis and multivariate regression analysis). (e-h) Caudate SBR was positively associated with BC of left parahippocampal gyrus, DC of left M1, nodal local efficiency of right rolandic operculum, and nodal efficiency of left M1 (FDR-corrected *p* < 0.05 in both Pearson correlation analysis and multivariate regression analysis). (i-l) Putamen SBR was positively associated with BC of left parahippocampal gyrus, DC of left M1, nodal local efficiency of right rolandic operculum, and nodal efficiency of left M1 (FDR-corrected *p* < 0.05 in both Pearson correlation analysis and multivariate regression analysis). The association analysis between graphical network metrics and SBR levels was conducted by Pearson correlation method and multivariate regression analysis with age, sex, disease duration, and years of education as covariates. FDR-corrected *p* < 0.05 was considered statistically significant. Abbreviations: BC, Betweenness centrality; DC, Degree centrality; SBR, Striatal binding ratio.

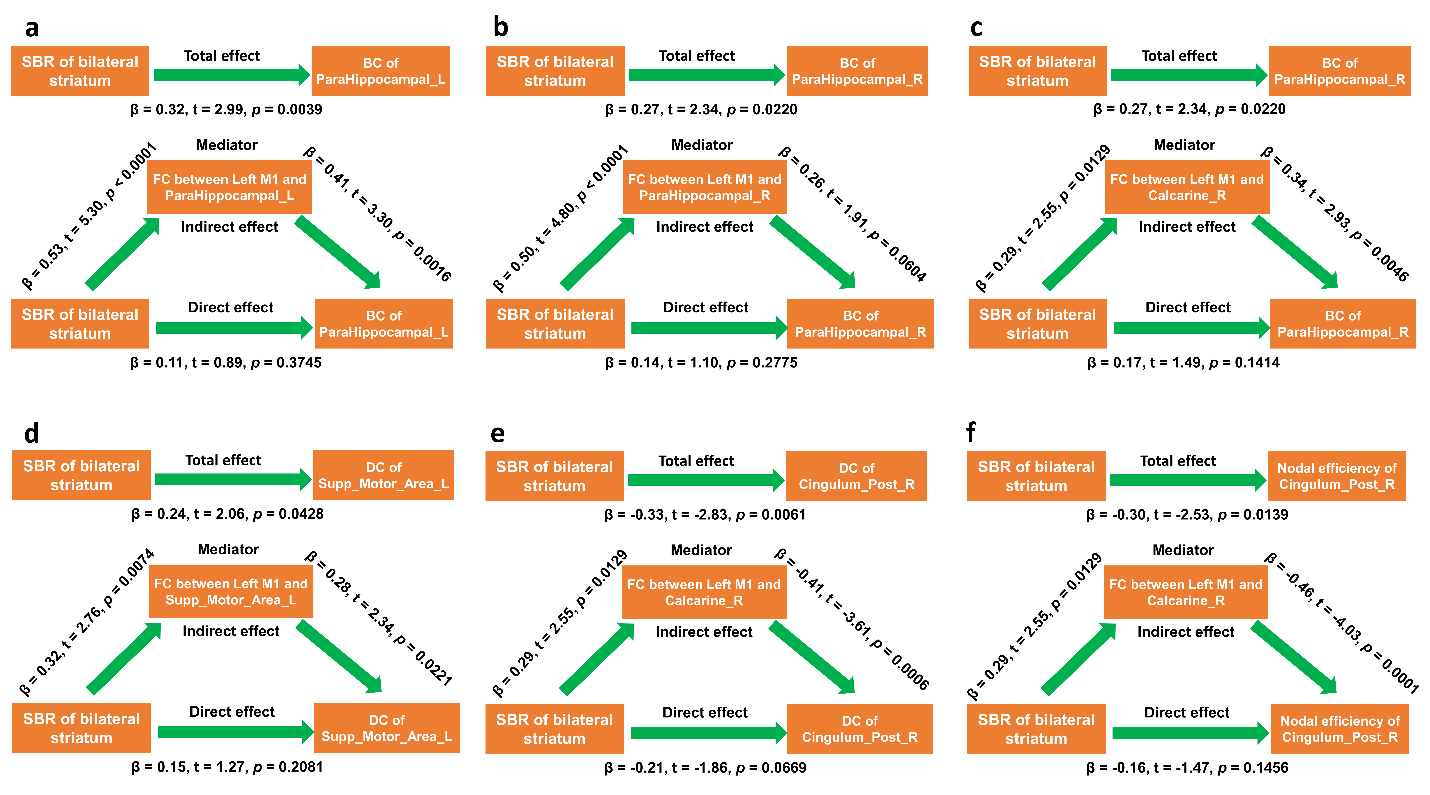

**Supplementary Figure 4. FC of left M1 mediated the effects of striatal dopamine depletion on other topological metrics.** (a) FC between left M1 and left parahippocampal gyrus mediated the effects of striatum SBR on BC of left parahippocampal gyrus. (b) FC between left M1 and right parahippocampal gyrus mediated the effects of striatum SBR on BC of right parahippocampal gyrus. (c) FC between left M1 and right calcarine mediated the effects of striatum SBR on BC of right parahippocampal gyrus. (d) FC between left M1 and left SMA mediated the effects of striatum SBR on DC of left SMA. (e) FC between left M1 and right calcarine mediated the effects of striatum SBR on DC of right posterior cingulate cortex. (f) FC between left M1 and right calcarine mediated the effects of striatum SBR on nodal efficiency of right posterior cingulate cortex. Mediation analysis was performed with age, sex, disease duration, and years of education as confounding variables. *p* < 0.05 was considered statistically significant. Abbreviations: BC, Betweenness centrality; DC, Degree centrality; M1, Primary motor cortex; FC, Functional connectivity; SBR, Striatal binding ratio; SMA, Supplementary motor area.

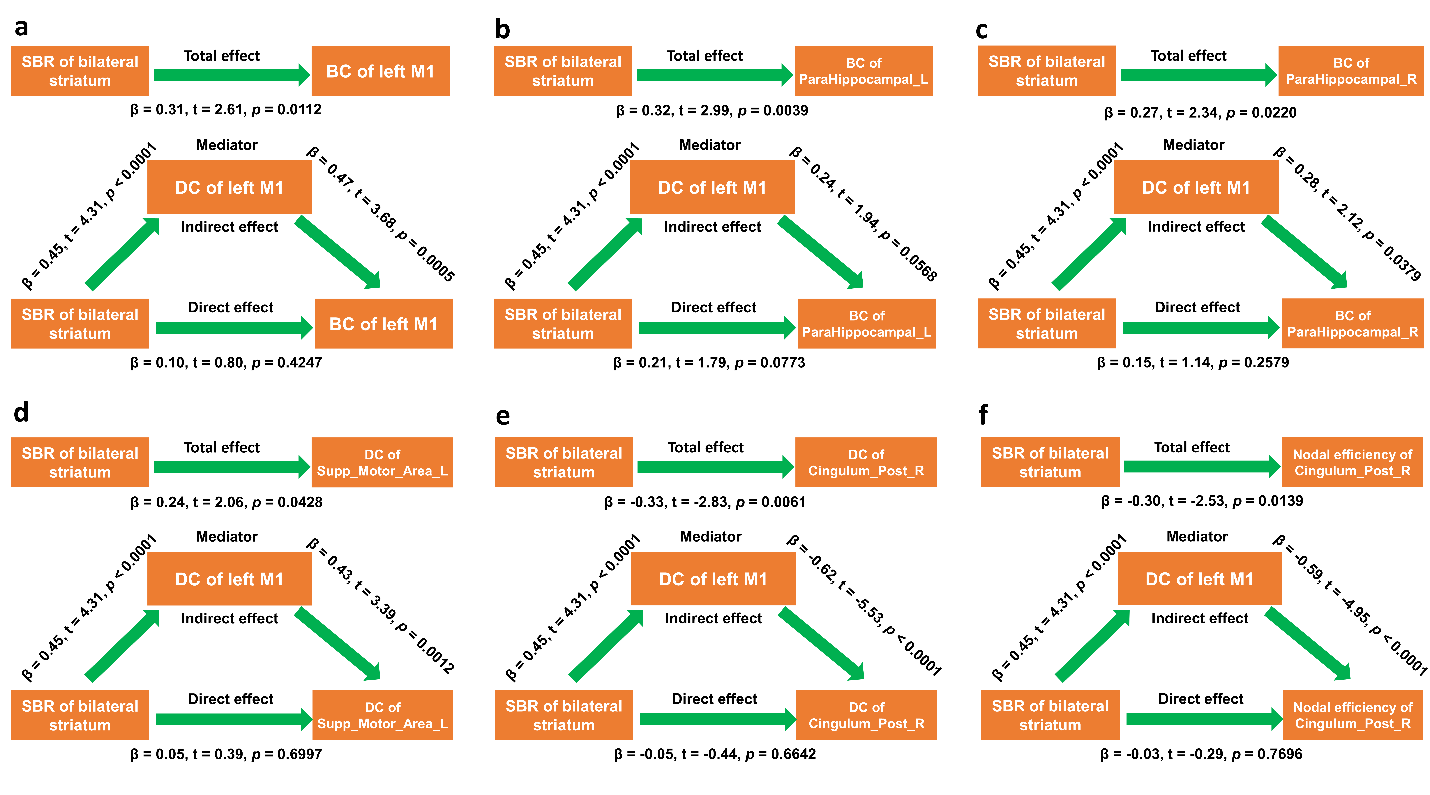

**Supplementary Figure 5. DC of left M1 mediated the effects of striatal dopamine depletion on topological metrics.** (a) DC of left M1 mediated the effects of striatum SBR on BC of left M1. (b) DC of left M1 mediated the effects of striatum SBR on BC of left parahippocampal gyrus. (c) DC of left M1 mediated the effects of striatum SBR on BC of right parahippocampal gyrus. (d) DC of left M1 mediated the effects of striatum SBR on DC of left SMA. (e) DC of left M1 mediated the effects of striatum SBR on DC of right posterior cingulate cortex. (f) DC of left M1 mediated the effects of striatum SBR on nodal efficiency of right posterior cingulate cortex. Mediation analysis was performed with age, sex, disease duration, and years of education as confounding variables. *p* < 0.05 was considered statistically significant. Abbreviations: BC, Betweenness centrality; DC, Degree centrality; M1, Primary motor cortex; SBR, Striatal binding ratio; SMA, Supplementary motor area.

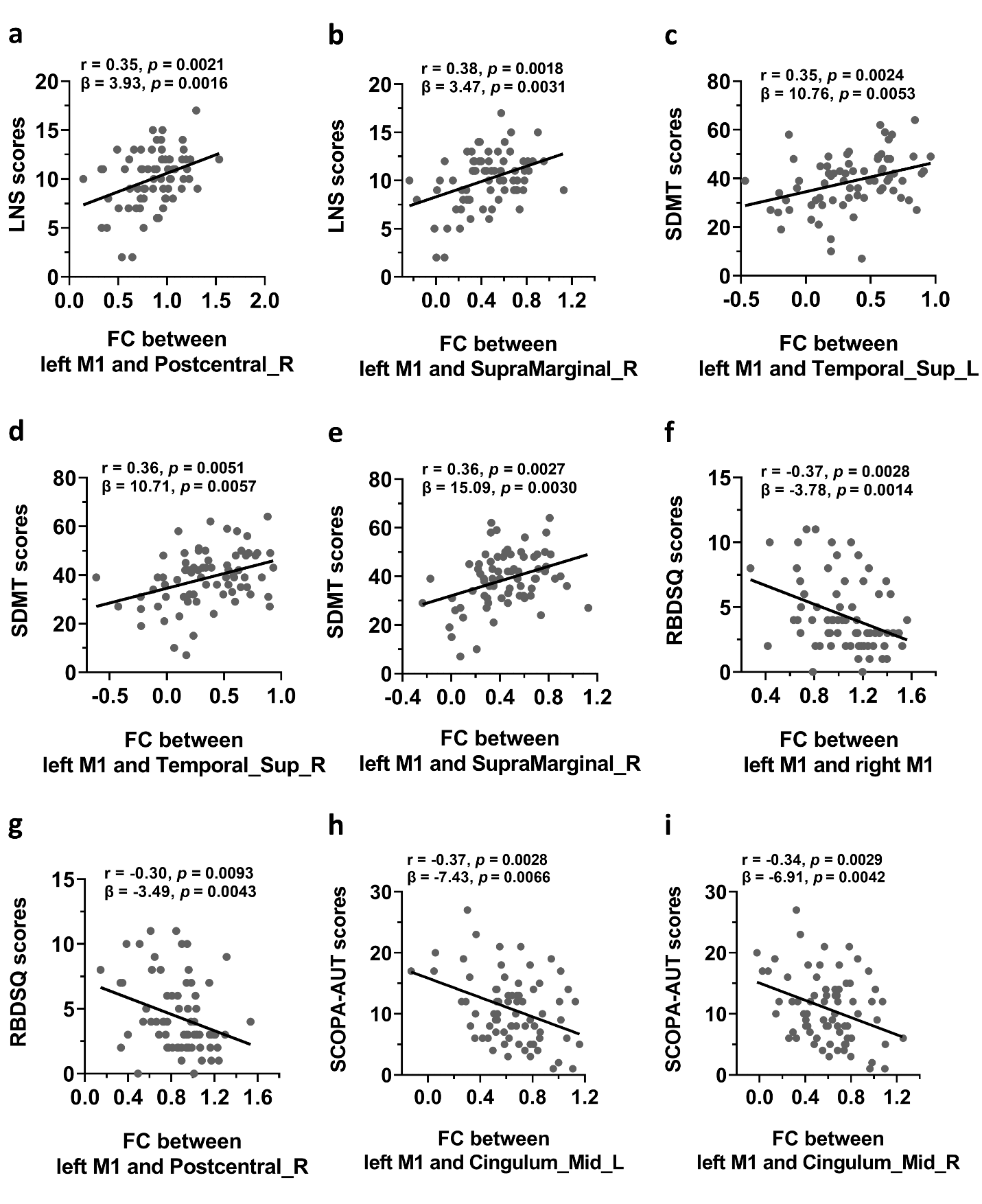

**Supplementary Figure 6. The FC of left M1 was significantly associated with non-motor symptoms.** (a-b) The FC between left M1 and right postcentral gyrus and right supramarginal gyrus were positively associated with LNS scores (FDR-corrected *p* < 0.05). (c-e) The FC between left M1 and left superior temporal gyrus, right superior temporal gyrus, right supramarginal gyrus were positively associated with SDMT scores (FDR-corrected *p* < 0.05). (f-g) The FC between left M1 and right M1 and right postcentral gyrus were negatively associated with RBDSQ scores (FDR-corrected *p* < 0.05). (h-i) The FC between left M1 and left middle cingulate gyrus and right middle cingulate gyrus were negatively associated with SCOPA-AUT scores (FDR-corrected *p* < 0.05). The association analysis between FC of left M1 and non-motor assessments was conducted by Pearson correlation method and multivariate regression analysis with age, sex, disease duration, and years of education as covariates. FDR-corrected *p* < 0.05 was considered statistically significant. Abbreviations: RBDSQ, REM Sleep Behavior Disorder Screening Questionnaire; SCOPA-AUT, Scale for Outcomes in Parkinson's Disease-Autonomic; LNS, Letter Number Sequencing test; SDMT, Symbol Digit Modalities Test; FC, Functional connectivity; M1, Primary motor cortex.

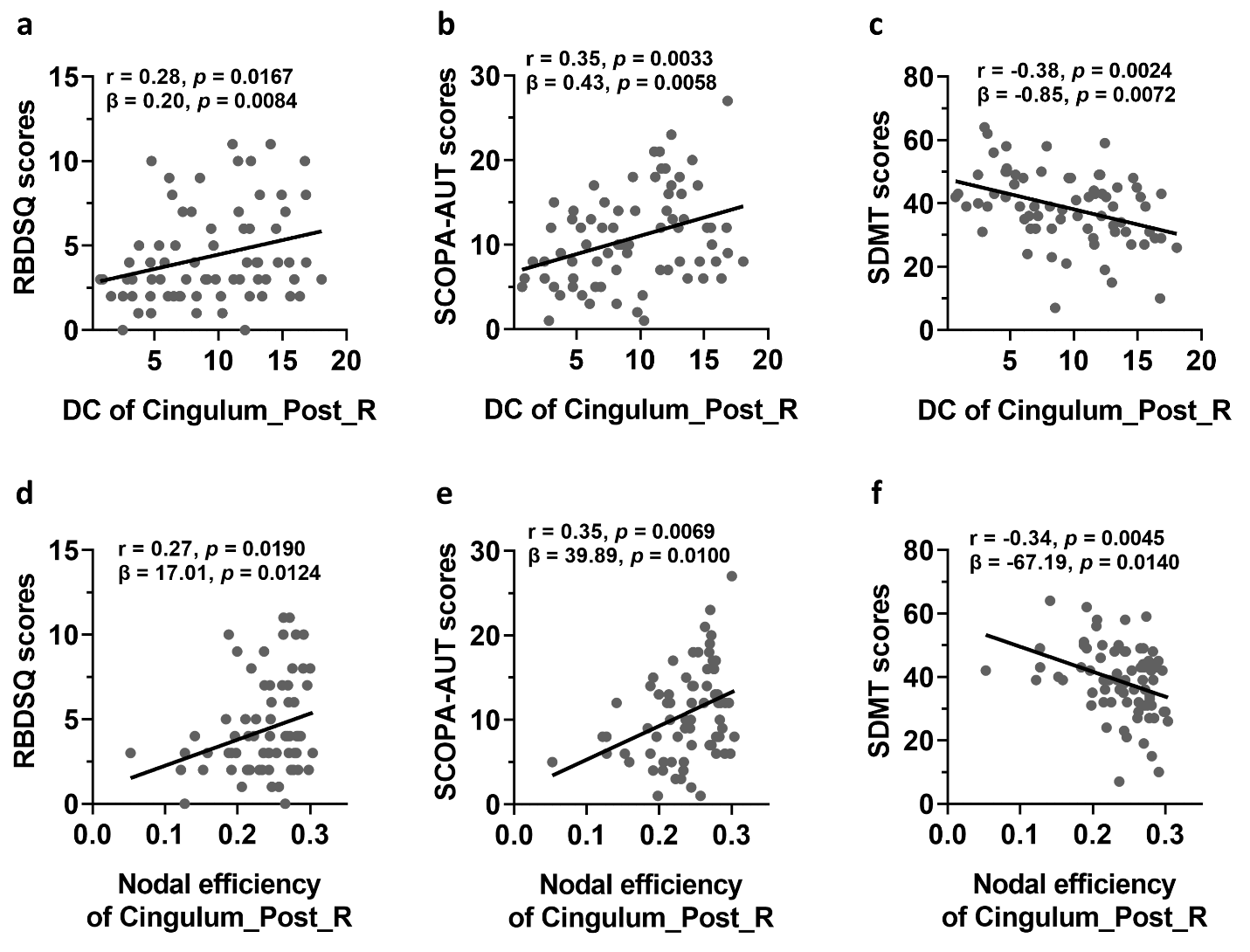

**Supplementary Figure 7. Associations between clinical variables and nodal network metrics of functional network.** (a-c) DC of right posterior cingulate cortex was significantly associated with scores of RBDSQ, SCOPA-AUT, and SDMT (FDR-corrected *p* < 0.05 in both Pearson correlation analysis and multivariate regression analysis). (d-f) Nodal efficiency of right posterior cingulate cortex was significantly associated with scores of RBDSQ, SCOPA-AUT, and SDMT (FDR-corrected *p* < 0.05 in both Pearson correlation analysis and multivariate regression analysis). The association analysis between graphical network metrics and clinical variables was conducted by Pearson correlation method and multivariate regression analysis with age, sex, disease duration, and years of education as covariates. FDR-corrected *p* < 0.05 was considered statistically significant. Abbreviations: DC, Degree centrality; RBDSQ, REM Sleep Behavior Disorder Screening Questionnaire; SCOPA-AUT, Scale for Outcomes in Parkinson's Disease-Autonomic; SDMT, Symbol Digit Modalities Test.

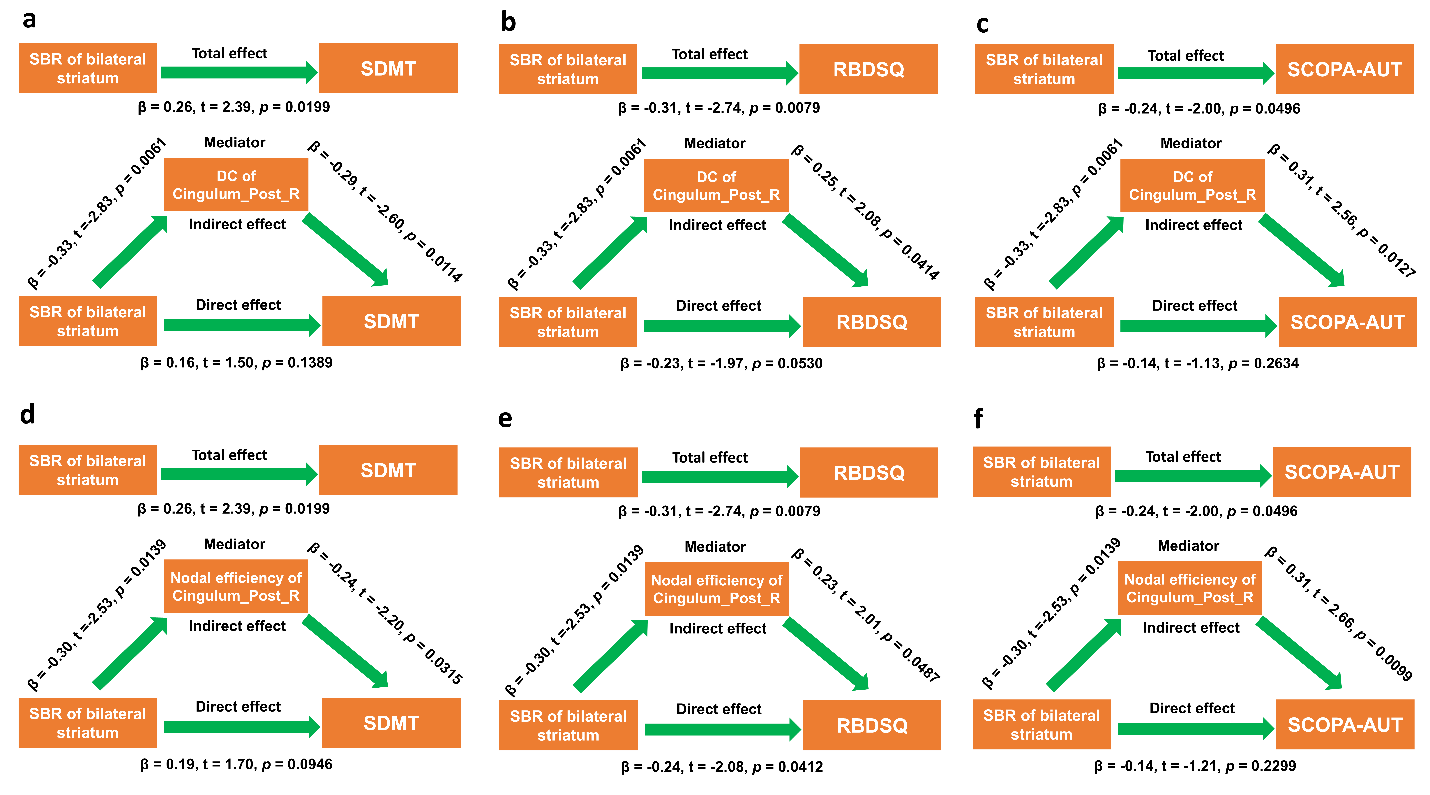

**Supplementary Figure 8. Nodal metrics of right posterior cingulate cortex mediated the effects of striatum SBR on non-motor assessments.** (a-c) DC of right posterior cingulate cortex mediated the effects of striatum SBRs on the scores of SDMT (a), RBDSQ (b), and SCOPA-AUT (c). (d-f) Nodal efficiency of right posterior cingulate cortex mediated the effects of striatum SBRs on the scores of SDMT (d), RBDSQ (e), and SCOPA-AUT (f). Mediation analysis was performed with age, sex, disease duration, and years of education as confounding variables. *p* < 0.05 was considered statistically significant. Abbreviations: DC, Degree centrality; RBDSQ, REM Sleep Behavior Disorder Screening Questionnaire; SCOPA-AUT, Scale for Outcomes in Parkinson's Disease-Autonomic; SDMT, Symbol Digit Modalities Test; SBR, Striatal binding ratio.
